# Frequency of transmission, asymptomatic shedding, and airborne spread of *Streptococcus pyogenes* among schoolchildren exposed to scarlet fever: a longitudinal multi-cohort molecular epidemiology contact tracing study

**DOI:** 10.1101/2021.07.04.21259990

**Authors:** Rebecca Cordery, Amrit K. Purba, Lipi Begum, Ewurabena Mills, Mia Mosavie, Ana Vieira, Elita Jauneikaite, Rhoda CY Leung, Matthew K. Siggins, Derren Ready, Peter Hoffman, Theresa Lamagni, Shiranee Sriskandan

## Abstract

**Background:** Despite recommendations regarding prompt treatment of cases and enhanced hygiene measures, scarlet fever outbreaks increased in England between 2014-2018. We aimed to assess the impact of standard interventions on transmission of *Streptococcus pyogenes* to classroom contacts, households, and classroom environments.

**Methods:** We undertook a prospective, contact tracing study in schools with 2 consecutive scarlet fever cases that were reported to local Health Protection teams in London between March 1^st^ and May 31st in 2018 and 2019. We cultured throat swabs from cases, household contacts, and classroom contacts at four time points. We also cultured hand swabs and cough plates from all cases, and from classroom contacts in 2019. Surface swabs from toys and other fomites in classrooms were cultured in 2018, and settle plates from classrooms were collected in 2019. Any sample with *S. pyogenes* detected was recorded as positive and underwent *emm* genotyping and genome sequencing to compare with the outbreak strain.

**Findings:** Six classes, comprising 12 scarlet fever cases, 17 household contacts, and 278 classroom contacts were recruited from March 1^st^ to May 31st in 2018 and the same period in 2019. Prevalence of the outbreak *S. pyogenes* strain in throat swabs from asymptomatic classroom contacts was high, increasing from 9.6% (11/115) in week 1, to 26.9% (34/126) in week 2, to 24.1% (26/108) in week 3, then 14.3% (5/35) in week 4. Colonisation with non-outbreak and non-genotyped *S. pyogenes* strains was 1.7% (2/115); 3.9% (5/126); 5.6% (6/108); and 0/35 in the same weeks. Genome sequencing showed clonality of isolates within each of six classes, confirming recent transmission accounted for high carriage. When transmissibility was tested, children who were asymptomatic carriers of *emm*4 and *emm*3.93 had positive cough plates on 1/14 (7.1%) and 7/21 (33.3%) occasions respectively. Only 1/60 surface swabs taken in 3 classrooms yielded the outbreak *S. pyogenes* strain. In contrast, 2/12 and 6/12 settle plates placed in elevated locations yielded the outbreak *S. pyogenes* strain in the two classrooms tested.

**Interpretation:** *S. pyogenes* transmission in schools is intense and may occur prior to, or inspite of reported treatment of cases, underlining a need for rapid case management. Despite guideline adherence, heavy shedding of *S. pyogenes* by small numbers of classroom contacts may perpetuate outbreaks, and airborne transmission has a plausible role in spread. The findings highlight the need for research to improve understanding and assess effectiveness of interventions to reduce *S. pyogenes* airborne transmission.

## Introduction

Since 2014, England has experienced rates of scarlet fever that are unprecedented in modern times (1, 2). Scarlet fever is a highly communicable exanthem caused by *Streptococcus pyogenes* that predominantly affects younger children. Over 30,000 cases were notified in 2018, the highest number since 1960, with an age-specific incidence of 523 per 100,000 among 1-4 year olds in England and Wales (3). Infections have long-been recognised to seed outbreaks in schools and nurseries, creating a significant public health burden (2). Though benign if treated promptly, there is a 20-fold increased risk of invasive streptococcal infection in households that include a recent case of scarlet fever (4). In 2016, a near 2-fold increase in invasive disease notifications was accompanied by emergence of a new sublineage of *emm*1 *S. pyogenes* (M1_UK_) expressing increased amounts of scarlet fever toxin (5).

The literature highlights a role for fomite surfaces in propagation of *S. pyogenes* outbreaks (6,7). Guidance for management of school/nursery outbreaks of scarlet fever emphasises the importance of maintaining good hand hygiene, prompt antibiotic treatment of index cases, and exclusion from school until 24h after antibiotic treatment has started (8). Escalation of interventions including daily cleaning of toys, utensils, and frequent touch points is recommended if necessary, along with daily vacuuming of soft furnishings (8).

To understand why outbreaks might continue to occur despite following these interventions, we investigated *S. pyogenes* transmission during the scarlet fever seasons of 2018 and 2019, focussing on educational settings with two sequential notifications of scarlet fever. Our primary aim was to determine if existing guidance that focuses on hygiene, alongside case exclusion, prevented transmission of *S. pyogenes* in the classroom. Our secondary aim was to investigate modes of transmission, to inform future guidance and provide insight into *S. pyogenes* outbreaks more generally.

## Methods

### Study design and participants

We conducted a prospective observational study in Greater London with recruitment from March 1 to May 31 2018 (Year 1), and March 1 to May 31 2019 (Year 2).

Schools and nurseries from the Greater London area that were notified to local Health Protection teams were invited to participate if they had two confirmed or probable scarlet fever cases aged 2-8 years from the same class within ten days of each other, with the most recent case arising in the preceding 48h. (Appendix p2 and p19). Routine public health advice was provided including advice that cases should be excluded from school until the case had received at least 24h of antibiotic treatment (8). Based on a pragmatic approach, the first location with 2 eligible cases that agreed to participate each month was selected.

### Procedures

Confirmed and probable cases in the affected class were invited to participate in daily sampling as soon as possible after diagnosis and up to day 8 of treatment, followed by samples collected at weekly intervals for 3 more weeks, comprising throat swabs, hand swabs, and Columbia blood agar (CBA, Oxoid, Basingstoke, UK) cough plates (appendix p19). Cases were visited at home during initial periods of school exclusion.

All household and classroom contacts of each case were invited to participate in weekly throat swab sampling for 4 weeks, or a total of 4 occasions if the weekly cycle was interrupted by holidays. In Year 2, classroom contacts were additionally asked to provide hand swabs and cough plates. To compare prevalence of carriage during an outbreak with prevalence after an outbreak, the protocol also incorporated a ‘break’ in the sampling schedule in Year 2 such that the 4^th^ sampling visit occurred 2-3 months after the 3^rd^ visit, with flexibility to adjust this period to remain within the school term. Any child with clinical pharyngitis or scarlet fever was directed to their primary care physician for management; swab results were not reported to participants or their physicians.

Routine advice on cleaning (8) was issued prior to environmental sampling. Environmental swabs were obtained from 20 frequently-touched items including toys and equipment in each classroom in 2018 in week 1. In 2019, to sample classroom air, CBA settle plates were positioned in each of two classrooms at heights of ∼1.5-2m for 2-3h on a weekly basis (Appendix p19-20).

After overnight culture, DNA was extracted from any *S. pyogenes* identified and *emm* genotyping performed, followed by genome sequencing, as detailed (Appendix p20). Genome sequences (Appendix p9-13) were compared with *S. pyogenes* sequences from an earlier survey of isolates associated with scarlet fever (9) (Appendix p14, p19-21); *emm*1 and *emm*4 lineages were assigned as reported (5, 10). Genome sequencing data are available from European Nucleotide Archive using the reference PRJEB43915. To evaluate sensitivity of conventional culture, DNA extracted from culture-negative swabs from one setting was subject to *S. pyogenes* ProS PCR (Appendix p19).

The study protocol was approved by a Research Ethics Committee (reference 18/LO/0025; IRAS 225006). Written informed consent was provided by parents or guardians, and assent provided by each child.

### Statistical analysis

Descriptive statistics only were used to analyse the data.

## Results

During the study, 156 educational outbreaks in London were reported in year 1 (2018), and 47 in year 2 (2019). Six settings were included, and 12 cases were recruited (Figure 1, Appendix, p3). Two cases were recruited per setting in settings 1-3; setting 2 and 3 were different classes in the same school. Three cases were recruited from each of settings 4 and 6. Cases declined to participate in setting 5, although contact and environment testing was undertaken based on one confirmed and one probable case. Sampling was completed by June 7^th^ in year 1 and June 28^th^ in year 2. During the study, participating schools reported increased prevalence of pharyngitis in addition to the reported cases of scarlet fever. The median attack rate for confirmed and probable cases was 5% (IQR 4.1-12.6%) and 10.1% (IQR 7.3-24.9%) with inclusion of possible cases. (Appendix, p3)

**Figure 1.**
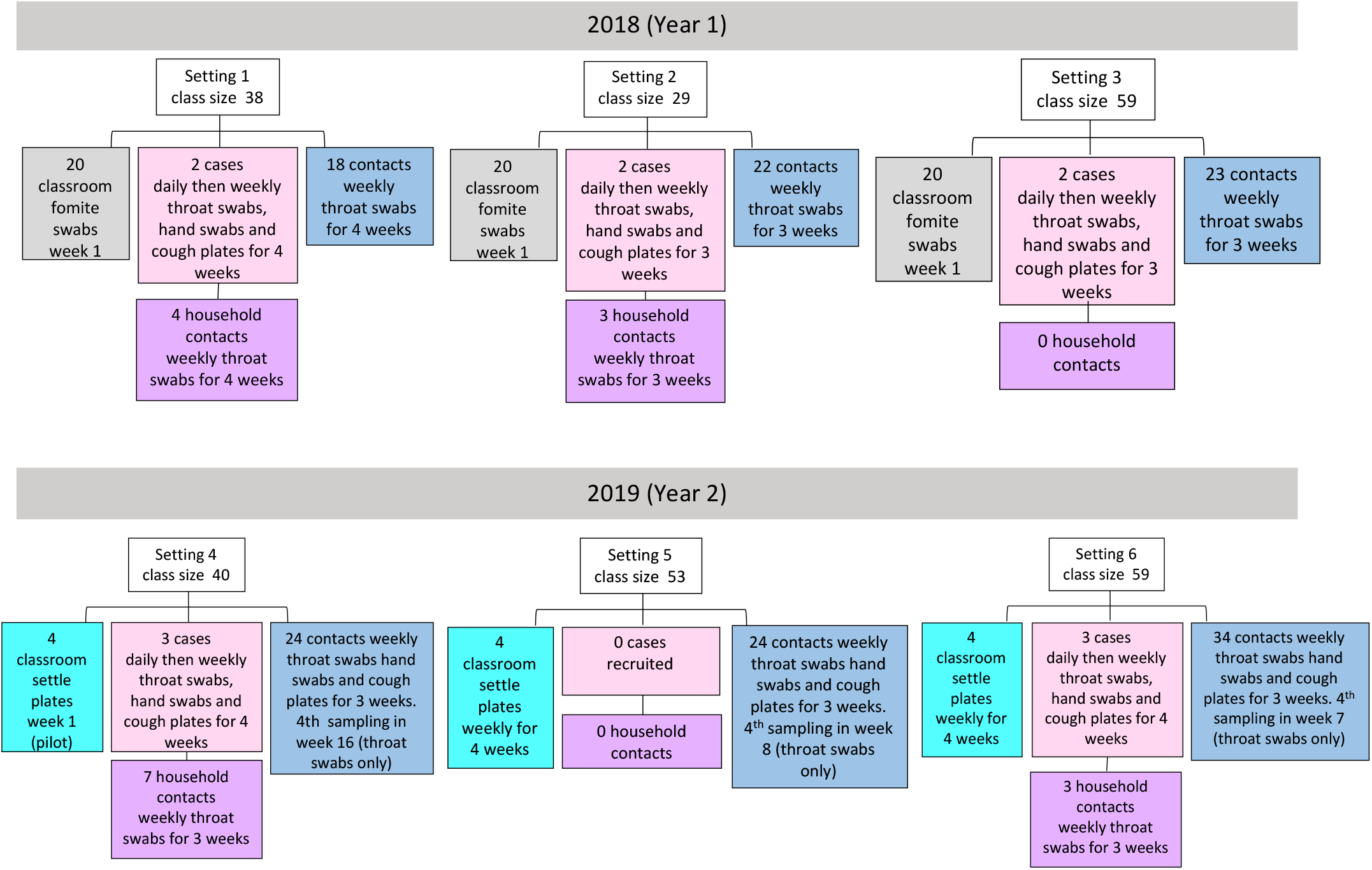
Flowchart indicating sample types sought from participants and schools in each setting. Samples provided, timings, and any breaks in sampling schedule are listed in Appendix pages 4-6.

Of the scarlet fever cases, 6/12 had been swabbed by primary care physicians prior to antibiotic treatment. *S. pyogenes* genotypes associated with each outbreak were derived from the swabs obtained from these cases, or inferred from the *S. pyogenes* isolates cultured from household contacts of cases. Genotypes were *emm*6, *emm*1, *emm*4, and *emm*3.93 (Figure 2). *Emm*1 isolates all belonged to the newly described M1_UK_ lineage (5), while *emm*4 isolates all belonged to the M4-complete lineage (10).

**Figure 2.**
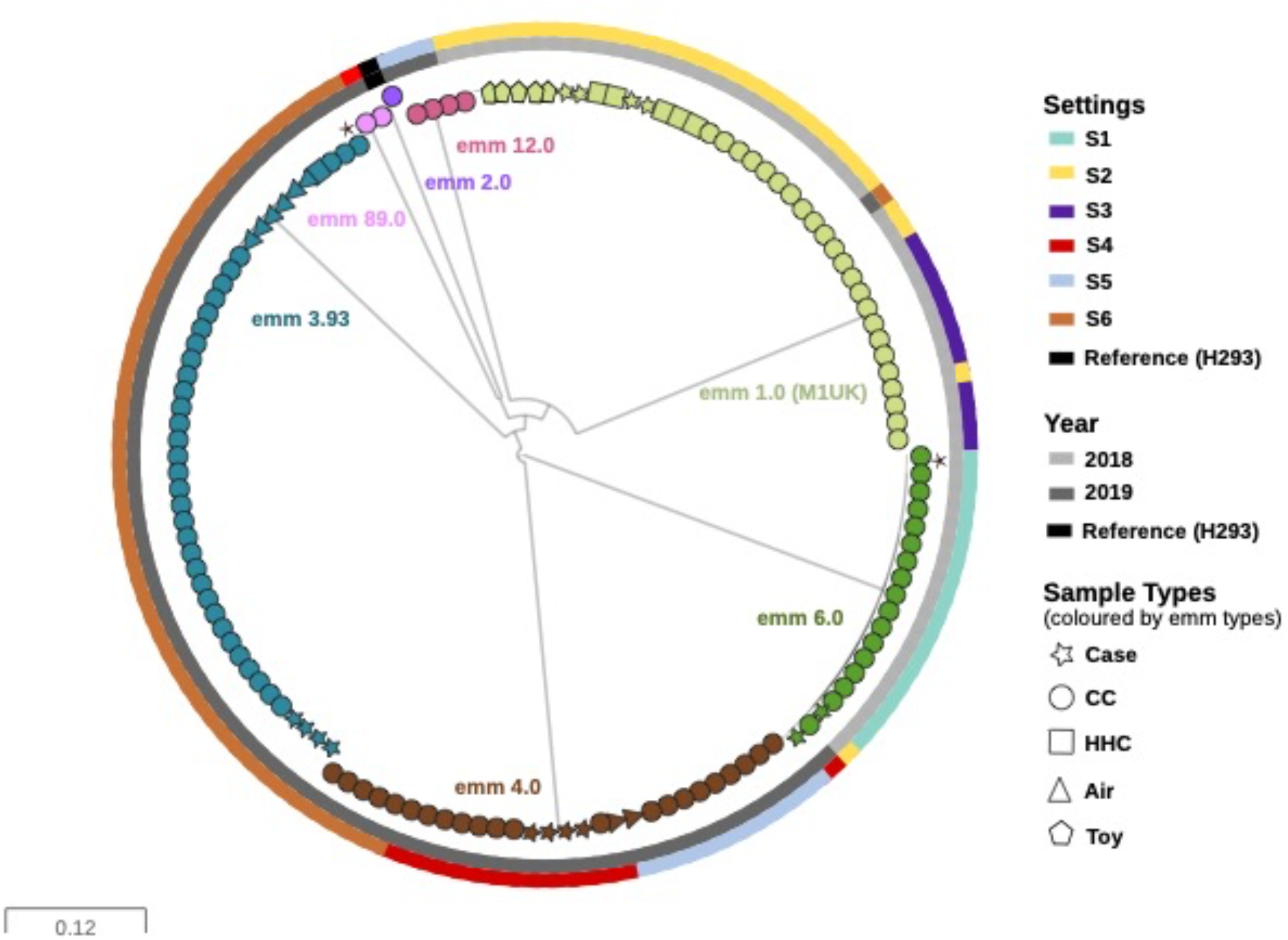
Phylogenetic relationship between *S. pyogenes* isolates sequenced from cases, contacts and environment in each of six settings. Maximum likelihood phylogenetic tree constructed from 20,229 core SNPs (without recombination regions) extracted after mapping 136 *S. pyogenes* isolates to the reference sequence H293 (*emm*89, HG316453.2). The outer rings (from outermost to innermost) represent the settings (S1-S6), and the year of collection. The tree was drawn in a circular layout with tips coloured based on *emm* types. The shape of the tip indicates the source (sample type) of individual isolates: Case; Classroom contact (CC); household contact (HHC); and environmental samples from settle plate (Air); or fomite (Toy). In some cases and contacts, multiple isolates were detected per participant (Appendix pages 4-6 and 9-13). Uncommon strain subtypes (*emm*3.143 and *emm*6.9) within an *emm*-type are indicated by *. For detailed analysis of each setting, see supplementary figures.

All bar one of the 12 cases received antibiotic treatment prescribed by the primary care physician. *S. pyogenes* was not detected in swab samples obtained during the first week, in all cases where antibiotic treatment had been initiated prior to the study visit (Table 1). The child who did not receive antibiotic treatment remained throat swab positive up to and including week 2 but had cleared *emm*4 *S. pyogenes* by week 3 (Table 1, appendix p4). One contact who had a positive throat swab and cough plate on day 1 of week 1 with *emm*3.93 *S. pyogenes*, developed scarlet fever the same day, so was also recruited as a case. Where the duration of antibiotic treatment was known (8/12 cases), 4 children completed a 10-day course, and 4 took an incomplete course (including 2 cases where a 7-day course was prescribed). Of those who had received antibiotic treatment, including those who completed a full 10-day course, the throat swab grew the prevailing outbreak strain of *S. pyogenes* again in 4/11 cases (36.3%); by week 2 (one with *emm*3.93), and by week 3 (two with M1_UK_ and one with *emm*3.93). One of the cases, carrying M1_UK,_ was also cough-plate and hand-swab positive in week 3 despite previously testing negative. (Table 1 and Appendix p4).

**Table 1.** *S. pyogenes* carriage and shedding in twelve cases of scarlet fever after recruitment to study.

| Positive swabs | Pre-study<sup>#</sup> | Week 1<sup>¶</sup> |  |  | Week 2 | Week 3 | Week 4 | Week 7-8 | Week 16 |
| --- | --- | --- | --- | --- | --- | --- | --- | --- | --- |
|  |  | Day 1 | Day 2 | Day 3 |  |  |  |  |  |
| Antibiotic<sup>§</sup> | 10/12 | 10/12 | 11/12 | 11/12 | 9/12 | 0/12 | 0/12 | 0/12 | 0/12 |
| Throat positive | 5/6 | 2/12 | 1/6 | 1/5 | 2/11 | 4/10 | 0/5 | 1/3 | 1/3 |
| Cough positive | - | 1/12 | 0/6 | 0/5 | 0/11 | 1/10 | 0/5 | - | - |
| Hand positive | - | 0/12 | 0/6 | 0/5 | 0/11 | 1/10 | 0/5 | - | - |

<sup>#</sup>Only 6 cases were swabbed by primary care physician pre-study.
<sup>¶</sup>Children were swabbed daily in week 1, until at least 72 h antibiotic therapy was given. Day 1 indicates first day of study and first swab.
<sup>§</sup>One case was identified in a contact at start of study so started antibiotics after first swab. One case received no antibiotics.
Final week of swabbing was week 4 (5 cases); week 7 (3 cases) or week 16 (3 cases).

Seventeen household contacts were enrolled in the study, representing 9 households (**Figure 1**). Of these, 3/17 (17.6%) had throat swabs that yielded *S. pyogenes*, two of whom had symptomatic pharyngitis with M1_UK_ and both demonstrated carriage spanning 2 or 3 weeks. (Appendix p5)

Among all classroom contacts swabbed in schools (Figure 1), asymptomatic throat carriage of the genomically-defined outbreak *S. pyogenes* strains among swabbed children rose from 11/115 (9.6%) in week 1, to 34/126 (26.9%) in week 2, to 26/108 (24.1%) in week 3, then 5/35 (14.3%) in week 4. This fell to 6/53 (11.3%), then 0/18 (0%) in classes where delayed swabbing was undertaken (Table 2). In contrast to carriage of the outbreak strain, median carriage of non-outbreak *S. pyogenes* was 2.8% (IQR 0-6.61%) during the study (Table 2). Transmission appeared particularly intense in setting 2, where 8/18 (44.4%) of children swabbed were infected with the outbreak M1_UK_ strain in week 2, in addition to the two household contacts (Figure 3, appendix p6). Overall, across all six settings, more than half of eligible children participated in the study (median participation rate 52.5%, IQR 43.71-65.83%; appendix p6).

**Table 2.**
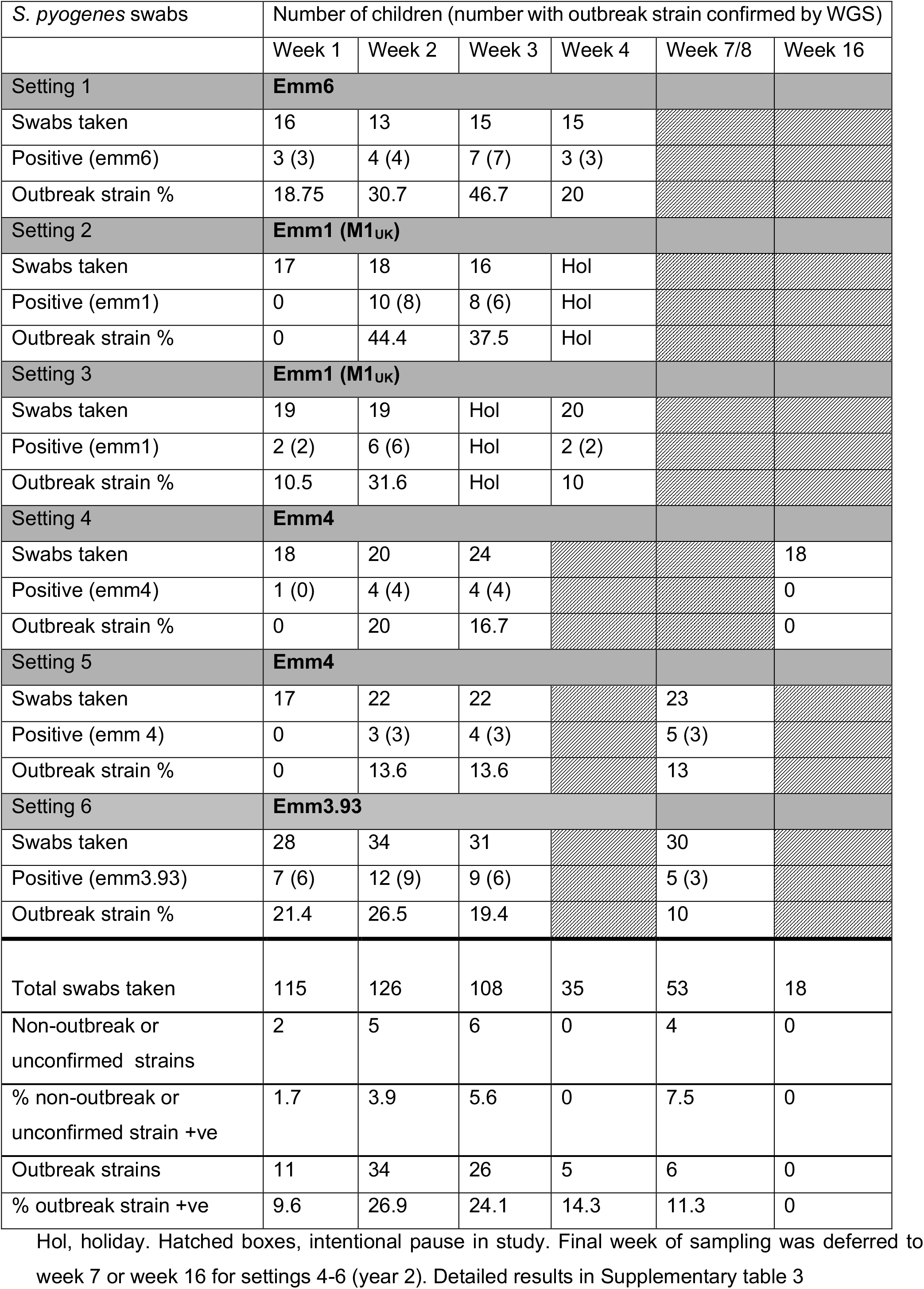
*S. pyogenes* outbreak strain throat swab prevalence in classroom contacts.

| *S. pyogenes* swabs | Number of children (number with outbreak strain confirmed by WGS) |  |  |  |  |  |
| --- | --- | --- | --- | --- | --- | --- |
|  | Week 1 | Week 2 | Week 3 | Week 4 | Week 7/8 | Week 16 |
| **Setting 1** | **Emm6** |  |  |  |  |  |
| Swabs taken | 16 | 13 | 15 | 15 |  |  |
| Positive (emm6) | 3 (3) | 4 (4) | 7 (7) | 3 (3) |  |  |
| Outbreak strain % | 18.75 | 30.7 | 46.7 | 20 |  |  |
| **Setting 2** | **Emm1 (M1<sub>UK</sub>)** |  |  |  |  |  |
| Swabs taken | 17 | 18 | 16 | Hol |  |  |
| Positive (emm1) | 0 | 10 (8) | 8 (6) | Hol |  |  |
| Outbreak strain % | 0 | 44.4 | 37.5 | Hol |  |  |
| **Setting 3** | **Emm1 (M1<sub>UK</sub>)** |  |  |  |  |  |
| Swabs taken | 19 | 19 | Hol | 20 |  |  |
| Positive (emm1) | 2 (2) | 6 (6) | Hol | 2 (2) |  |  |
| Outbreak strain % | 10.5 | 31.6 | Hol | 10 |  |  |
| **Setting 4** | **Emm4** |  |  |  |  |  |
| Swabs taken | 18 | 20 | 24 |  |  | 18 |
| Positive (emm4) | 1 (0) | 4 (4) | 4 (4) |  |  | 0 |
| Outbreak strain % | 0 | 20 | 16.7 |  |  | 0 |
| **Setting 5** | **Emm4** |  |  |  |  |  |
| Swabs taken | 17 | 22 | 22 |  | 23 |  |
| Positive (emm 4) | 0 | 3 (3) | 4 (3) |  | 5 (3) |  |
| Outbreak strain % | 0 | 13.6 | 13.6 |  | 13 |  |
| **Setting 6** | **Emm3.93** |  |  |  |  |  |
| Swabs taken | 28 | 34 | 31 |  | 30 |  |
| Positive (emm3.93) | 7 (6) | 12 (9) | 9 (6) |  | 5 (3) |  |
| Outbreak strain % | 21.4 | 26.5 | 19.4 |  | 10 |  |
| **Total swabs taken** | **115** | **126** | **108** | **35** | **53** | **18** |
| Non-outbreak or unconfirmed strains | 2 | 5 | 6 | 0 | 4 | 0 |
| % non-outbreak or unconfirmed strain +ve | 1.7 | 3.9 | 5.6 | 0 | 7.5 | 0 |
| Outbreak strains | 11 | 34 | 26 | 5 | 6 | 0 |
| % outbreak strain +ve | 9.6 | 26.9 | 24.1 | 14.3 | 11.3 | 0 |

Hol, holiday. Hatched boxes, intentional pause in study. Final week of sampling was deferred to week 7 or week 16 for settings 4-6 (year 2). Detailed results in Supplementary table 3

**Figure 3.**
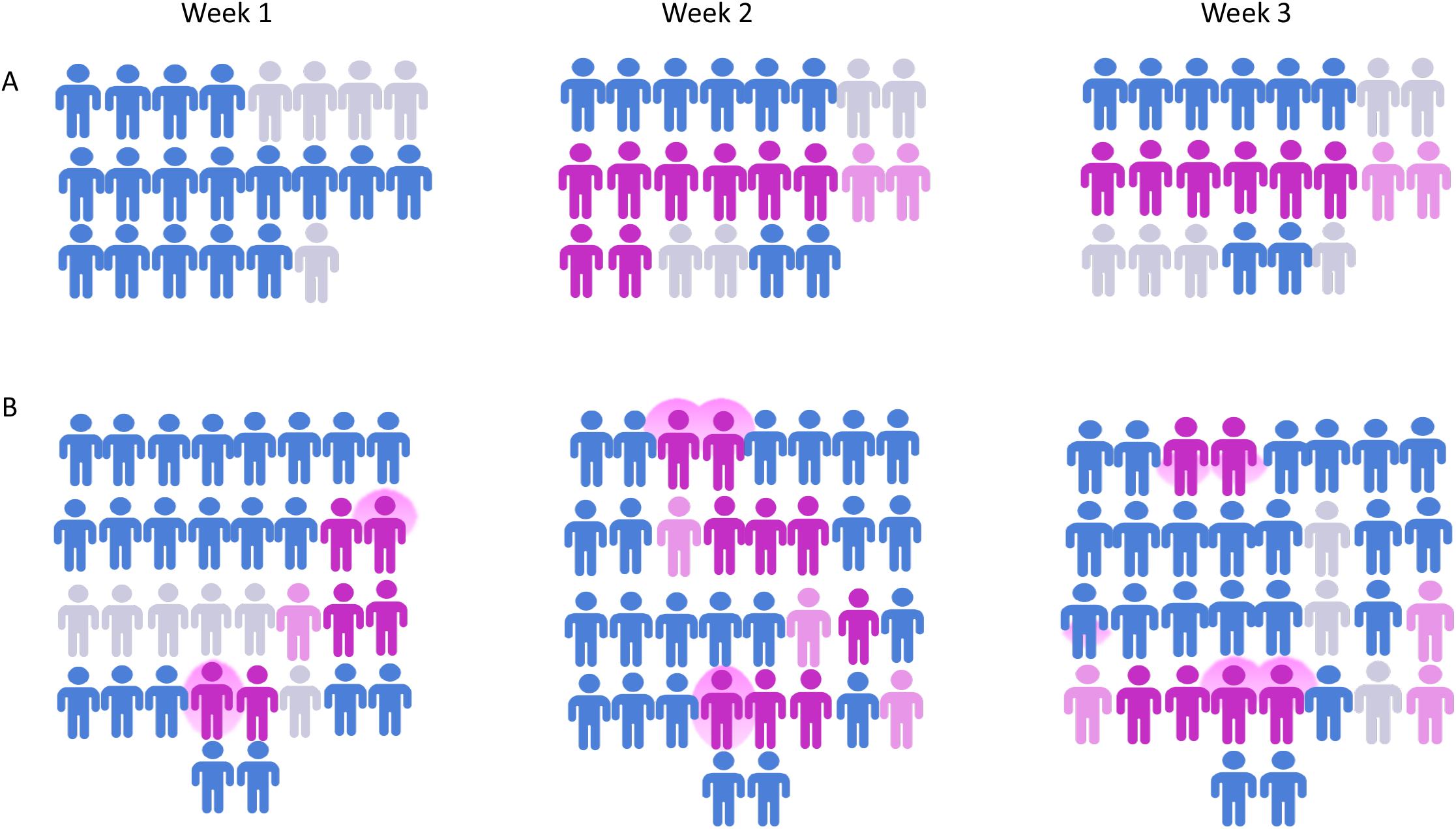
Acquisition of M1_UK_ and *emm*3.93 *S. pyogenes* by classroom contacts. Classroom contacts in (A) setting 2 and (B) Setting 6. Classroom contacts had throat swabs weekly for at least three weeks after identification of case; each icon represents an individual child, grouped by week of swabbing. Colour of icons (adioma.com) indicates throat swab result: Blue, negative; dark pink, outbreak strain *S. pyogenes*; pale pink, non-outbreak strain or genotype not confirmed; grey, participant not swabbed in that week. Pink glow behind icon indicates cough plate and/or hand swab positive. For (A) (n=22 classroom contacts) *S. pyogenes* was identified in 10/18 (55.5%) and 8/16 (50.0%) in weeks 2 and 3 respectively. The outbreak strain (*emm*1/M1_UK_) was identified in 8/18 (44.4%) and 6/16 (37.5%) in weeks 2 and 3. For (B) (n=34 classroom contacts) *S. pyogenes* was identified in 7/28 (25.0%), 12/34 (35.2%), and 9/30 (30.0%) in weeks 1, 2, and 3 respectively. The outbreak strain *emm*3.93 was identified in 6/28 (21.4%), 9/34 (26.5%), and 6/30 (20.0%) of children who had swabs taken in weeks 1, 2, and 3 respectively. Fourth week not shown.

Whole genome sequencing was undertaken on all viable *S. pyogenes* isolates identified. Within each setting, isolates of the same genotype were clonal, with individual strains being a median of 0 SNPs (range 0-5 SNPs) different to other strains in that setting, consistent with a common source of transmission, but a median of 12 SNPs (range 4-55 SNPs) different from the closest strain of the same genotype sequenced in 2014. Core genomes exhibited more similarity than expected. (Appendix p8, p15-18).

Environmental swabs taken in settings 1-3 in week 1 of the first year of the study yielded mixed bacterial growth (∼1 × 10^2^ cfu – 1 × 10^4^ cfu per swab; appendix p7). *S. pyogenes* was identified from just one toy (train track) from setting 2: Five colonies/swab of the outbreak strain *emm*1 (M1_UK_) were 1-3 SNPs different to isolates obtained from children in the same class (appendix p16).

To determine the source of ongoing transmission in the classrooms, the protocol was amended in year 2 to include hand-swabs and cough-plates from classroom contacts in addition to throat swabs, while settle plates were used to sample air in the classroom. In setting 4, where *emm*4 predominated, one of four children carrying *S. pyogenes* was cough-plate positive in the second week, but not in subsequent weeks (appendix p6); all isolates in this setting were 0 SNP apart (appendix p17). Cough-plates were negative in setting 5 (also *emm*4 where again isolates were 0 SNP apart; appendix p6, p17). Overall 1/14 (7.1%) *emm*4 positive throat swabs was associated with a positive cough plate. In setting 6, where *emm*3.93 *S. pyogenes* predominated, cough-plates were positive for the outbreak *emm*3.93 strain in one third of children with positive *emm*3.93 throat swabs (week 1, 2/6; week 2, 3/9, week 3, 2/6) (Figure 3 and Appendix p6). The vast majority of strains were 0 SNP different from one another, bar three strains that were 1, 2, and 5 SNP apart (Appendix p18). SNPs differentiating strains within each outbreak are listed (Appendix p8).

In setting 4, where prevalence of the outbreak *emm*4 *S. pyogenes* had been 0/18(0%), 4/20(20%), and 4/24(16.6%) in weeks 1-3 respectively, final samples from classroom contacts were taken in week 16 after an intentional break in sampling, and all (0/18) were negative. It was only possible to incorporate a 4-week gap for settings 5 and 6 for logistic reasons related to the timing of the school term. In setting 5, carriage of the outbreak *emm*4 strain remained steady throughout the study, with a prevalence of 0% (0/17), 13.6% (3/22), 13.6% (3/22) in weeks 1-3 respectively and 12.5% (3/23) at week 7. Notably 2/3 of *emm*4 carriers who were positive in week 7 had also been positive in week 3. In setting 6, where prevalence of the outbreak *emm*3.93 strain in weeks 1, 2, and 3 was 21.4% (6/28), 26.5% (9/34), 19.4% (6/31) (Figure 3) respectively in asymptomatic classroom contacts, carriage of *emm*3.93 fell to 10% (3/30) by week 8. Notably in this outbreak, throat swabs from two of the cases and a single household contact were positive in week 3, *emm3*.93 (Appendix p4-5). To determine the likelihood that culture-based sampling might be insensitive, DNA extracted from culture-negative swabs from setting 6 was subject to *S. pyogenes* ProS PCR; 50/65 (76.9%) culture negative swabs were deemed qPCR negative.

Four settle plates were used per classroom in settings 5 and 6 to sample room air, sited above child head height. In setting 5, one of four settle plates (placed above a whiteboard) was positive for *emm*4 *S. pyogenes* in each of weeks 2 and 3 of the study (Table 3). In setting 6, two of four settle plates were positive for *emm*3.93 *S. pyogenes* in weeks 1, 2, and 3 of the study, including plates on top of 2m cupboards (Table 3). In both settings, *S. pyogenes* strains on settle plates were identical to strains identified in children (*emm*4 and *emm*3.93, appendix p17-18).

**Table 3.**
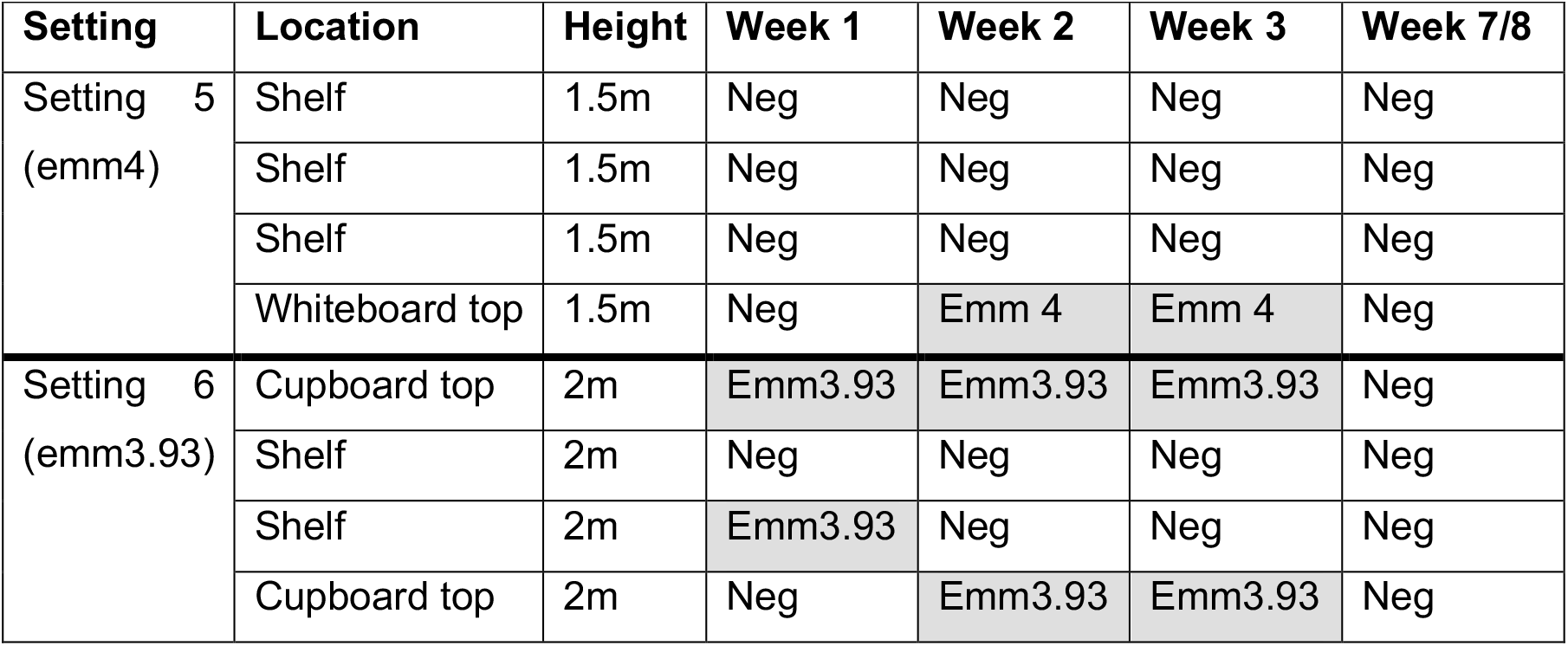
Air settle plate results from settings 5 and 6.

| **Setting** | **Location** | **Height** | **Week 1** | **Week 2** | **Week 3** | **Week 7/8** |
| --- | --- | --- | --- | --- | --- | --- |
| Setting 5<br>(emm4) | Shelf | 1.5m | Neg | Neg | Neg | Neg |
|  | Shelf | 1.5m | Neg | Neg | Neg | Neg |
|  | Shelf | 1.5m | Neg | Neg | Neg | Neg |
|  | Whiteboard top | 1.5m | Neg | Emm 4 | Emm 4 | Neg |
| Setting 6<br>(emm3.93) | Cupboard top | 2m | Emm3.93 | Emm3.93 | Emm3.93 | Neg |
|  | Shelf | 2m | Neg | Neg | Neg | Neg |
|  | Shelf | 2m | Emm3.93 | Neg | Neg | Neg |
|  | Cupboard top | 2m | Neg | Emm3.93 | Emm3.93 | Neg |

## Discussion

This prospective contact tracing study of *S. pyogenes* transmission was undertaken in response to an unprecedented rise in scarlet fever notifications in England. Using genome sequencing to confirm common sources of transmission, we found a high prevalence of the outbreak strain among asymptomatic classroom contacts, peaking in the second week of our investigations. Despite antibiotic treatment and isolation of index cases for 24h after initiation of antibiotic treatment, and implementation of standard hygiene measures within the classrooms, transmission within the class was observed. Enhanced sampling in year 2 revealed evidence of prominent *S. pyogenes* shedding by some children, and airborne dispersal of genomically identical strains in the classroom.

In the six settings investigated, *emm*1 (M1_UK_) strains were involved in two outbreaks, *emm*4 in two outbreaks, and *emm*3.93 and *emm*6 strains were involved in one outbreak each. New scarlet fever-causing lineages have been associated with national upsurges in invasive *S. pyogenes* (5, 11), exemplified by emergence of M1_UK,_ a lineage that expresses increased levels of the scarlet fever toxin SpeA (5). There are however international differences in approach to treatment of streptococcal pharyngitis (12), and transmission risks are not widely considered (13). Coupled with the public health impact of outbreaks (2), high attack rates in schools (14), and increased risk of invasive infections (4), there is a rationale to limit the spread of *S. pyogenes* in the population, particularly where lineages such as *emm*1 and *emm*3, that are independently associated with high case fatality or severe manifestations (15), are involved.

Current public health guidance on scarlet fever management focusses on treatment, exclusion, and hygiene interventions, with escalation to include daily cleaning of surfaces (8). Although contamination of fomites including toys is no doubt important, we found that transmission was ongoing despite such cleaning and in the absence of frequent surface *S. pyogenes* contamination. Indeed, the main source of *S. pyogenes* appeared to be the children themselves. Other than foodborne scarlet fever outbreaks (16), there are remarkably few recent investigations that examine transmission routes. Four contemporary reports describe scarlet fever outbreaks with high attack rates (23-72%) in educational settings that were not controlled by standard interventions (17-20); transmission routes were investigated in just one (17). Three describe use of antibiotic treatment to end the outbreaks, treating all colonised children (17-19). Microbiological detection of *S. pyogenes* to enable treatment of children found to be colonised would however be challenging to undertake routinely for all scarlet fever outbreaks. Our study underlines a need for research to evaluate a potential role for molecular point of care tests for rapid detection of colonisation and outbreak management.

By week 3, prevalence of the genomically-confirmed outbreak strain was 14%-47% across all six settings, while carriage of non-outbreak strains was infrequent. Carriage rates detected are remarkably similar to data from higher attack rate outbreaks (17-20) and studies from the pre-antibiotic era, where 26-33% of streptococcal pharyngitis classroom contacts were observed to carry the outbreak serotype (21). Our study draws attention to the importance of context when measuring *S. pyogenes* carriage rates as, outside of the springtime outbreak season, asymptomatic carriage in healthy children in England is estimated at <6% (22) and <1% in healthy adults (23). Studies that report average annual rates of asymptomatic carriage therefore fail to recognise the very major impact of seasonal variation and outbreaks and may provide misleading contextual information when providing recommendations.

We frequently recovered the outbreak strain among classroom contacts in week 1 of sampling, indicating that transmission had already occurred in the classroom, whether from one of the index cases of scarlet fever or another unknown source. It is possible that treatment may have been delayed in some cases; a recent study identified one fifth of scarlet fever cases to be initially diagnosed as a viral infection, allowing transmission to continue if the affected child remains in school, untreated (24). Whether this is sufficient to explain scarlet fever outbreaks merits further analysis and modelling.

It is generally believed that asymptomatic carriage is unlikely to lead to transmission, although the term ‘carrier’ is more often used to refer to a case that has been treated for *S. pyogenes* pharyngitis but demonstrates microbiological failure (25). In our study, the systematic increase in prevalence of outbreak strains between weeks 1 and 2 in classroom contacts pointed to ongoing transmission from asymptomatic carriers, despite index cases having been excluded and treated. Indeed, after returning to school, four cases re-acquired the outbreak strain by week 3, presumably through contact with other colonised contacts or classroom air, although did not develop symptomatic illness; these cases re-acquired *emm*1 (M1_UK_, 2 cases) and *emm*3.93 (2 cases). Our investigations using cough plates and hand swabs demonstrated that at least one third of asymptomatic *emm*3.93 carriers were shedding more *S. pyogenes* than other children in exhaled cough and on hands. These findings raise a question about the definition of colonisation and infection in young children, who may not be able to identify their symptoms, or may be infectious without symptoms. The study confirms the existence of so-called heavy shedders among those who are apparently asymptomatic at the time of sampling and resonates with historic reports of heavy nasal shedding by *S. pyogenes* nasal carriers (26).

Settle plates placed in elevated locations for just 2-3 hours, provided evidence of *S. pyogenes* dispersal in classroom air in settings 5 and 6; the timing of settle plate positivity coincided with more intense periods of asymptomatic shedding. Settle plates provide an easy read out in an outbreak setting and could be used to indicate a need for improved ventilation, social distancing, or surveillance for heavy shedders. Airborne spread of scarlet fever-causing streptococci was previously recognised as a threat in hospitals in the 1930’s and in military barracks in the 1940’s (28). Contemporary outbreaks of *S. pyogenes* that are not explained by direct contact alone suggest a need to take not only indirect contact but also airborne transmission into consideration when developing guidelines.

The scarlet fever outbreak setting has provided an unexpected model for understanding *S. pyogenes* transmission and immunity. Children of the same age were exposed to a presumed similar inoculum of *S. pyogenes* in class, yet only ∼5% developed scarlet fever, some of whom re-acquired the outbreak strain without illness three weeks later, while some anecdotally developed pharyngitis. Others were found to asymptomatically shed *S. pyogenes* for several weeks, while others showed transient colonisation, or no infection whatsoever. Our understanding of immunity to *S. pyogenes* is heavily dominated by factors that influence susceptibility to invasive disease. The findings highlight a gap in our knowledge regarding mucosal immunity to *S. pyogenes*, specifically whether full immunity requires antibodies to prevent streptococcal adherence, promote bacterial clearance, and inhibit streptococcal virulence factors during pharyngitis. This is potentially critical to vaccine development. Coupled with differences in genetic susceptibility and oral microbiota, differing layers of immunity could go some way to explain why children express a range of disease phenotypes.

There are limitations to our study. Sampling of fomites was limited to single timepoints in year 1, while settle plates to sample air were only used in year 2, restricting ability to determine the relative importance of each transmission route. Our study was based in London, so findings may not be relevant to rural, or low- and middle-income settings, or regions with different climate. Furthermore, transmission intensity may be seasonal; we attempted to sample outside of the main scarlet fever season, but schools were unwilling to participate. Indeed, engagement was high when anxiety about scarlet fever was greatest.

The study has demonstrated that heavy asymptomatic shedding by a proportion of children may account for persistence of *S. pyogenes* outbreaks in classroom settings. The phenomenon of super-shedders and super-spreaders is increasingly recognised as a source of heterogeneity in infectious disease modelling (29, 30) and is consistent with the explosive nature of some streptococcal outbreaks. Relevance to other respiratory infections is unclear. Our findings may explain the periodic failure of interventions focussed on hygiene alone to curtail outbreaks of *S. pyogenes*, both in the classroom and other institutional settings. The recognition of heavy asymptomatic shedding highlights a potential role for social distancing, improved respiratory hygiene, and better ventilation in reducing *S. pyogenes* transmission during outbreaks. As an unforeseen consequence of the COVID-19 pandemic, implementation of these measures has proven highly successful in halting England’s scarlet fever upsurge, at least in 2020 (31), highlighting a need for better understanding of transmission routes in preventing future upsurges.

## Data Availability

All data pertaining to the study is included in the supplementary files and genomics data is available via the European Nucleotide Archive, under the accession number PRJEB43915

## Acknowledgements

The authors would like to acknowledge the support of the schools, nurseries, and staff, as well as the many children and households who participated in this study, and the support of London Health Protection Teams in PHE. The NIHR BRC Colebrook laboratory and BRC Genomics facility are also acknowledged. EJ is a Rosetrees/Stoneygate 2017 Imperial College Research Fellow, funded by Rosetrees Trust and the Stoneygate Trust (Fellowship no. M683).

## Author contributions

Conceptualisation and funding acquisition SS, RC, TL; Project supervision and methodology SS; Recruitment supervision RC; data collection LB, AKP, MM, EM, MKS, RCYL, DR, PH; data analysis AV, EJ; data visualisation RC, SS, AV, EJ; LB, AKP, and RC verified the identifiable participant data; EM, EJ, and AV verified linked non-identifiable microbiological and genomic data.

Original draft RC, SS; Review and editing AKP, EJ, AV, PH, DR, RCYL, TL; Final manuscript all authors. All authors had full access to all non-identifiable data in the study and agreed with the decision to submit for publication.

## Data sharing

All data are included in the supplementary materials or are available from European Nucleotide Archive using the reference PRJEB43915.

