## Supplementary Appendix for "Frequency of transmission, asymptomatic shedding, and airborne spread of *Streptococcus pyogenes* among schoolchildren exposed to scarlet fever: a longitudinal multi-cohort molecular epidemiology contact tracing study"

#### Contents

| | | Page |
|---|---|---|
| Supplementary Table 1. | Case Definitions | 2 |
| Supplementary Table 2. | Study Site Recruitment | 3 |
| Supplementary Table 3. | Cases in each setting- detailed results | 4 |
| Supplementary Table 4. | Household contacts in each setting- detailed results | 5 |
| Supplementary Table 5. | School contacts in each setting- detailed results | 6 |
| Supplementary Table 6. | Surface swab results (settings 1-3, 2018) | 7 |
| Supplementary Table 7. | SNPs between isolates within each cluster | 8 |
| Supplementary Table 8. | Accession numbers of genomes from each setting | 9-13 |
| Supplementary Table 9. | Genomes from Chalker et al (ref.12) used for analysis | 14 |
| Supplementary Figure S1. | Phylogenetic relationship of *emm*6 strains (setting 1). | 15 |
| Supplementary Figure S2. | Phylogenetic relationship of *emm*1 strains (settings 2 & 3). | 16 |
| Supplementary Figure S3. | Phylogenetic relationship of *emm*4 strains (settings 4 & 5). | 17 |
| Supplementary Figure S4. | Phylogenetic relationship of *emm*3 strains (setting 6). | 18 |
| Supplementary Methods | | 19-21 |
| Suppl. References | | 21 |

**Supplementary Table 1. Case Definitions**

| **Definitions** | |
|---|---|
| Confirmed | Clinical diagnosis of Scarlet Fever by a health professional and *S. pyogenes* detected on a throat swab |
| Probable | Clinical diagnosis of Scarlet Fever by a health professional |
| Possible | Case reported by a reliable source (e.g. nursery manager, school secretary), presenting with signs and symptoms consistent with scarlet fever, and a close epidemiological link e.g. household contact of a confirmed case; or attending school where there is a confirmed scarlet fever outbreak<br>Cases reported by a health professional where scarlet fever is part of a differential diagnosis and other infections may be just as likely. |
| Carriage | Throat swab positive for *S. pyogenes* in asymptomatic individual |
| Outbreak | 2 or more cases of scarlet fever linked in time, place and person i.e. in children or staff member, within 10 days of each other in the same school or nursery class. |

Inclusion criteria for participating schools were two confirmed or probable scarlet fever cases aged 2-8 years from the same class within ten days of each other, with the most recent case arising in the preceding 48h. Cases were prospectively swabbed if they were confirmed or probable.

**Supplementary Table 2. Study Site case and household recruitment by setting**

| | Age group (years) | Inferred outbreak emm type | Final scarlet fever outbreak size | Cases recruited | Whole class size | Attack rate in setting* | Household contacts recruited |
|---|---|---|---|---|---|---|---|
| 1 | 3-4 | Emm6 | 2 confirmed | 2 | 38 | 5.2% | 4 |
| 2 | 5-6 | Emm1 ($M1_{UK}$) | 4 probable<br>9 possible | 2 | 29 | 4.5-14.8% | 3 |
| 3 | 4-5 | Emm1 ($M1_{UK}$) | | 2 | 59 | | 0 |
| 4 | 3-4 | Emm4 | 4 confirmed<br>4 probable<br>6 possible | 3 | 40 | 20-35% | 7 |
| 5 | 4-5 | Emm4 | 1 confirmed<br>1 probable<br>3 possible | 0 | 53 | 3.8-9.4% | 0 |
| 6 | 4-5 | Emm3.93 | 3 probable<br>3 possible | 3 | 59 | 5.0-10.1% | 3 |

*Attack rate range derived from (confirmed + probable) and (confirmed + probable +possible) cases.

### Supplementary Table 3. Cases in each setting- detailed results

| | | *S. pyogenes* swab results, no. of children (outbreak strain confirmed by WGS) | | | | | | | | |
|---|---|---|---|---|---|---|---|---|---|---|
| | GP swab Y/N (result) | Week 1 | | | | Week 2 | Week 3 | Week 4 | Week 7/8 | Week 16 |
| | | Day 1 | Day 2 | Day 3 | Day 4 | | | | | |
| Setting 1 | | | | | | | | | | |
| **Case 1** | Y (**+**) | | | | | | | | | |
| Throat | | NEG | NEG | NEG | NEG | NEG | NEG | NEG | | |
| Cough | | NEG | NEG | NEG | NEG | NEG | NEG | NEG | | |
| Hand | | NEG | NEG | NEG | NEG | NEG | NEG | NEG | | |
| **Case 2** | Y (**+**) | | | | | | | | | |
| Throat | | NEG | NEG | NEG | NEG | NEG | NEG | NEG | | |
| Cough | | NEG | NEG | NEG | NEG | NEG | NEG | NEG | | |
| Hand | | NEG | NEG | NEG | NEG | NEG | NEG | NEG | | |
| Setting 2 | | | | | | | | | | |
| **Case 1** | N | | | | | | | | | |
| Throat | | NEG | NEG | ND | ND | NEG | **+** | ND | | |
| Cough | | NEG | NEG | ND | ND | NEG | **+** | ND | | |
| Hand | | NEG | NEG | ND | ND | NEG | **+** | ND | | |
| **Case 2** | N | | | | | | | | | |
| Throat | | NEG | NEG | NEG | ND | NEG | **+** | ND | | |
| Cough | | NEG | NEG | NEG | ND | NEG | NEG | ND | | |
| Hand | | NEG | NEG | NEG | ND | NEG | NEG | ND | | |
| Setting 3 | | | | | | | | | | |
| **Case 1** | N | | | | | | | | | |
| Throat | | NEG | ND | ND | ND | NEG | ND | NEG | | |
| Cough | | NEG | ND | ND | ND | NEG | ND | NEG | | |
| Hand | | NEG | ND | ND | ND | NEG | ND | NEG | | |
| **Case 2** | N | | | | | | | | | |
| Throat | | NEG | ND | ND | ND | ND | ND | NEG | | |
| Cough | | NEG | ND | ND | ND | ND | ND | NEG | | |
| Hand | | NEG | ND | ND | ND | ND | ND | ND | | |
| Setting 4 | | | | | | | | | | |
| **Case 1** | Y (-) | | | | | | | | | |
| Throat | | **+** | **+** | **+** | ND | **+** | NEG | | | **+** |
| Cough | | NEG | NEG | NEG | ND | NEG | NEG | | | ND |
| Hand | | NEG | NEG | NEG | ND | NEG | NEG | | | ND |
| **Case 2** | Y (**+**) | | | | | | | | | |
| Throat | | NEG | NEG | NEG | ND | NEG | NEG | | | NEG |
| Cough | | NEG | NEG | NEG | ND | NEG | NEG | | | ND |
| Hand | | NEG | NEG | NEG | ND | NEG | NEG | | | ND |
| **Case 3*** | Y (**+**) | | | | | | | | | |
| Throat | | ND | ND | ND | ND | NEG | NEG | NEG | | NEG |
| Cough | | ND | ND | ND | ND | NEG | NEG | NEG | | ND |
| Hand | | ND | ND | ND | ND | NEG | NEG | NEG | | ND |
| Setting 6 | | | | | | | | | | |
| **Case 1** | N | | | | | | | | | |
| Throat | | NEG | ND | ND | ND | **+** | **+** | | **+** | |
| Cough | | NEG | ND | ND | ND | NEG | NEG | | ND | |
| Hand | | NEG | ND | ND | ND | NEG | NEG | | ND | |
| **Case 2** | Y (**+**) | | | | | | | | | |
| Throat | | NEG | ND | ND | ND | NEG | **+** | | NEG | |
| Cough | | NEG | ND | ND | ND | NEG | NEG | | ND | |
| Hand | | NEG | ND | ND | ND | NEG | NEG | | ND | |
| **Case 3**** | N | | | | | | | | | |
| Throat | | **+** | ND | ND | ND | NEG | NEG | | NEG | |
| Cough | | **+** | ND | ND | ND | NEG | NEG | | ND | |
| Hand | | NEG | ND | ND | ND | NEG | NEG | | ND | |

Positive *S. pyogenes* result indicated by '+'; negative by 'NEG'. Hatched cells indicate intentional pause in study
*Case 3 in setting 4 was studied weekly only; **Case 3 in setting 6 was identified on day 1 of the study (initially recruited as a contact). Abbreviations: Hol, school holiday (unable to sample); WGS, whole genome sequencing; GP, General practitioner (primary care physician); Y, yes; N, no; ND, not done. Setting 5, no cases recruited.

**Supplementary Table 4. Household contacts in each setting- detailed results**

| | | | Week 1 | Week 2 | Week 3 | Week 4 |
|---|---|---|---|---|---|---|
| Setting 1 emm6 | | | | | | |
| Case1 | HHC1 | | NEG | NEG | NEG | NEG |
| | HHC2 | | NEG | NEG | NEG | NEG |
| Case 2 | HHC1 | | NEG | NEG | NEG | ND |
| | HHC2 | | ND | NEG | NEG | ND |
| Setting 2 emm1 ($M1_{UK}$) | | | | | | |
| Case 1 | HHC1 | | **+** | **+** | NEG | |
| Case 2 | HHC1 | | NEG | NEG | NEG | |
| | HHC2 | | **+** | **+** | **+** | |
| Setting 4 emm4 | | | | | | |
| Case 1 | HHC1 | | NEG | NEG | NEG | |
| | HHC2 | | NEG | NEG | NEG | |
| | HHC3 | | NEG | NEG | ND | |
| Case 2 | HHC 1 | | NEG | NEG | NEG | |
| | HHC2 | | ND | ND | NEG | |
| Case 3 | HHC1 | | NEG | NEG | NEG | |
| | HHC2 | | NEG | NEG | NEG | |
| Setting 6 emm3.93 | | | | | | |
| Case 1 | HHC1 | | NEG | NEG | NEG | |
| | HHC2 | | NEG | NEG | **+** | |
| Case 2 | HHC1 | | NEG | NEG | ND | |
| Overall Household contacts | | | | | | |
| Participant total | | | 17 | 17 | 17 | 4 |
| Swabs taken | | | 15 | 16 | 16 | 2 |
| *S. pyogenes* positive (%) | | | 2 (13.3) | 2 (12.5) | 2 (12.5) | 0 (0) |
| Outbreak strain (%) | | | 2 (13.3) | 2 (12.5) | 2 (12.5) | 0 (0) |

Positive *S. pyogenes* result indicated by '+'; negative by 'NEG'.
Abbreviations. HHC, household contact.

**Supplementary Table 5. School contacts in each setting- detailed results**

| | *S. pyogenes* sample results, Number of children (outbreak strain confirmed by *emm* type or WGS) | | | | | | |
|---|---|---|---|---|---|---|---|
| | Week 1 | Week2 | Week 3 | Week 4 | Week 7/8 | Week 16 | Whole class size |
| Setting 1 | | | | | | | |
| TS Positive | 3 (3) | 4 (4) | 7 (7) | 3$^{\Psi}$ (3) | | | 38 |
| TS Negative | 13 | 9 | 8 | 12 | | | |
| TS swab total | 16 | 13 | 15 | 15 | | | |
| Absent/Ref | 2 | 5 | 3 | 3 | | | |
| Participant total | 18 | 18 | 18 | 18 | | | |
| Setting 2 | | | | | | | |
| TS Positive | 0 | 10 (8)$^{\Psi\Psi\Psi}$ | 8 (6) | Hol | | | 29 |
| TS Negative | 17 | 8 | 8 | Hol | | | |
| TS swab total | 17 | 18 | 16 | - | | | |
| Absent/Ref | 5 | 4 | 6 | Hol | | | |
| Participant total | 22 | 22 | 22 | Hol | | | |
| Setting 3 | | | | | | | |
| TS Positive | 2 (2) | 6 (6) | Hol | 2 (2) | | | 59 |
| TS Negative | 17 | 13 | Hol | 18 | | | |
| TS swab total | 19 | 19 | - | 20 | | | |
| Absent/Ref | 4 | 4 | Hol | 3 | | | |
| Participant total | 23 | 23 | Hol | 23 | | | |
| Setting 4 | | | | | | | |
| TS Positive | 1 (0) | 4 (4) | 4 (4) | | | 0 | 40 |
| TS Negative | 17 | 16 | 20 | | | 18 | |
| TS swab total | 18 | 20 | 24 | | | 18 | |
| Cough positive | 0 | 1 (1) | 0 | | | ND | |
| Hand positive | 0 | 0 | 0 | | | ND | |
| Absent/Ref | 0 | 4 | 0 | | | 7 | |
| Participant total | 18 | 24 | 24 | | | 25 | |
| Setting 5 | | | | | | | |
| TS Positive | 0 | 3 (3) | 4 (3) | | 5 (3) | | 53 |
| TS Negative | 17 | 19 | 18 | | 18 | | |
| TS swab total | 17 | 22 | 22 | | 23 | | |
| Cough positive | 0 | 0 | 0 | | ND | | |
| Hand positive | 0 | 0 | 0 | | ND | | |
| Absent/Ref | 1 | 1 | 2 | | 1 | | |
| Participant total | 18 | 23 | 24 | | 24 | | |
| Setting 6♣ | | | | | | | |
| TS Positive | 7$^{\Psi}$ (6) | 12$^{\Psi}$ (9) | 9$^{\Psi}$ (6) | | 5$^{\Psi\Psi}$ (3) | | 59 |
| TS Negative | 21 | 22 | 22 | | 25 | | |
| TS swab total | 28 | 34 | 31 | | 30 | | |
| Cough positive | 2 (2) | 3 (3) | 2 (2) | | ND | | |
| Hand positive | 1 (1) | 1 (1) | 3$^{\Psi}$ (2) | | ND | | |
| Absent/Ref | 2 | 0 | 4 | | 4 | | |
| Participant total | 30 | 34 | 34 | | 34 | | |

Hatched cells indicate intentional pause in study (Settings 4-6, a break between 3rd week and final week was incorporated.)

Hol, unable to sample as school holiday; TS, throat swab; ND, not done;

♣Setting 6; one contact in week 1 with positive throat swab and cough plate was subsequently diagnosed as a case and given antibiotics, with all samples subsequently negative and is shown for completeness; week 3 samples taken at end of week 2 due to school holiday.

$^{\Psi}$one, $^{\Psi\Psi}$two, or $^{\Psi\Psi\Psi}$three samples have no WGS.

**Supplementary Table 6. Surface swab results (settings 1-3, year 1)**

| Setting | Sample type | Colony count | Setting | Sample type | Colony count | Setting | Sample type | Colony count |
|---|---|---|---|---|---|---|---|---|
| 1 | Construction toy | 100 | 2 | Activity table | 350 | 3 | Snack table | 10,000 |
| | Construction toy | 200 | | Activity table | 450 | | Wooden stool | 1,980 |
| | Construction toy | 100 | | Wooden track* | 1000 | | Smartboard screen | 60 |
| | Construction toy | 100 | | Magnetic letter | 150 | | Book | 100 |
| | Construction toy | 150 | | Magnetic letter | <25 | | Magnifier handle | 340 |
| | Toy car | 300 | | Lego brick | 100 | | Ipad screen | 720 |
| | Toy car | 250 | | Smart screen | 100 | | Door handle | 60 |
| | Book | 400 | | Book | 200 | | Small bin | 400 |
| | Book | 200 | | Book | 500 | | Lego table | 320 |
| | Large toy car | 350 | | iPad | 950 | | Activity table | 1,600 |
| | Play surface | 100 | | iPad | 1000 | | Dry wipe pen | <20 |
| | Toy animal | 200 | | Chair | 200 | | Activity table | 100 |
| | Toy animal | 350 | | Jigsaw piece | 100 | | Paint table | 180 |
| | Book | 500 | | Toy truck | 250 | | Glue bottle | 420 |
| | Book | 400 | | Plastic scissors | 550 | | Book | 220 |
| | Construction parts | 180 | | Plastic animal | 200 | | Toy Screwdriver | 180 |
| | Construction parts | 200 | | Book | 100 | | Plastic apron | 460 |
| | Toy cooker | 320 | | Phonics game | 3000 | | Paint brush handle | 1,320 |
| | Toy shell | 340 | | Counting toy | 200 | | Toy animal | 400 |
| | Plastic money | 200 | | Glue stick | 200 | | Chair | 380 |

Colony counts reflect mixed flora. No beta haemolytic streptococci except sample '*' yielding 5 colonies of *S. pyogenes*

**Supplementary Table 7. SNPs between isolates within each cluster**

| Classes | Emm type | PosGene | ID | Gene Name | Gene Product | SNP | Residue | Type of mutation | Variant prediction |
|---|---|---|---|---|---|---|---|---|---|
| **Setting 1** | Emm6 | 155402 | M6_RS00980 | NA | deoxynucleoside kinase | 135T>C | Asp45Asp | Syn | Low |
| | | 910654 | M6_RS04595 | *guaA* | glutamine-hydrolyzing GMP synthase | 201C>T | Tyr67Tyr | Syn | Low |
| | | 1277861 | M6_RS06460 | *arcC* | carbamate kinase | 279T>C | Asn93Asn | Syn | Low |
| | | 1323939 | M6_RS06670 | NA | Glycoside hydrolase family 125 protein | 180C>T | Ser60Ser | Syn | Low |
| | | 1880406 | M6_RS09345 | *hasB* | UDP-glucose 6-dehydrogenase HasB | 172C>T | Gln58* | Stop | High |
| | | 1881063 | M6_RS09345 | *hasB* | UDP-glucose 6-dehydrogenase HasB | 829C>A | Gln277Lys | Non-Syn | Moderate |
| | | 1894721 | x | x | x | x | | Intragenic | x |
| **Setting 2** | Emm1 | 14547 | M5005_RS00060 | NA | ATP-dependent metallopeptidase FtsH/Yme1/Tma family protein | 1743T>C | Arg581Arg | Syn | Low |
| | | 907441 | M5005_RS04580 | *guaA* | glutamine-hydrolyzing GMP synthase | 1540C>T | Pro514Ser | Non-Syn | Moderate |
| | | 1011845 | M5005_RS05145 | *ssb* | Single-stranded DNA-binding protein | 215G>T | Gly72Val | Non-Syn | Moderate |
| | | 1477185 | M5005_RS07535 | NA | MptD family putative ECF transporter S Component | 448A>G | Thr150Ala | Non-Syn | Moderate |
| **Setting 3** | Emm1 | 43445 | M5005_RS00285 | *purN* | phosphoribosylglycinamide formyltransferase | 48C>A | Val16Va | Syn | Low |
| | | 940218 | M5005_RS04730 | NA | M1 family metallopeptidase | 1098A>G | Pro366Pro | Syn | Low |
| | | 1255526 | x | x | x | x | | intragenic | x |
| **Setting 4** | Emm4 | None | None | None | None | None | None | None | None |
| **Setting 5** | Emm4 | None | None | None | None | None | None | None | None |
| **Setting 6** | Emm3 | 1882546 | SpyM3_1852 | *hasB* | putative UDP-glucose 6-dehydrogenase | 754C>T | Pro252Ser | Non-Syn | Moderate |
| **Setting 4 and Setting 6** | Emm4 | 899513 | MGAS10750_Spy 0941 | NA | Xaa-His dipeptidase | 1268C>A | Thr423Lys | Non-Syn | Moderate |
| | | 1010790 | MGAS10750_Spy 1067 | NA | hypothetical protein | 913G>A | Asp305Asn | Non-Syn | Moderate |
| | | 1026739 | MGAS10750_Spy 1079 | NA | ATPase associated with chromosome architecture/replication | 121G>A | Ala41Thr | Non-Syn | Moderate |
| | | 1450398 | MGAS10750_Spy 1513 | NA | PTS system, galactose-specific IIB component | 62C>T | Ala21Val | Non-Syn | Moderate |
| | | 1735343 | x | x | x | x | | Intragenic | x |

**Supplementary Table 8 Accession numbers of genomes from each setting**

| Sample ID | Individual ID | Year | Setting | Source | Sample type | Timepoint (1-4) | emm typing | MLST | Accession* | Unique Name |
|---|---|---|---|---|---|---|---|---|---|---|
| A_C1_T_001 | A-C1 | 2019 | Setting 4 | Case | Throat swab | 1 | EMM4.0 | 39 | ERS6110953 | SAMEA8426050 |
| A_C1_T_002 | A-C1 | 2019 | Setting 4 | Case | Throat swab | 1 | EMM4.0 | 39 | ERS6110954 | SAMEA8426051 |
| A_C1_T_003 | A-C1 | 2019 | Setting 4 | Case | Throat swab | 1 | EMM4.0 | 39 | ERS6110955 | SAMEA8426052 |
| A_C1_T_008 | A-C1 | 2019 | Setting 4 | Case | Throat swab | 2 | EMM4.0 | 39 | ERS6110956 | SAMEA8426053 |
| A_C1_T_106 | A-C1 | 2019 | Setting 4 | Case | Throat swab | 4 | EMM6.0 | 382 | ERS6110957 | SAMEA8426054 |
| A_CC18_C_008 | A_CC18 | 2019 | Setting 4 | CC | Cough plate | 2 | EMM4.0 | 39 | ERS6110958 | SAMEA8426055 |
| A_CC18_T_008 | A_CC18 | 2019 | Setting 4 | CC | Throat swab | 2 | EMM4.0 | 39 | ERS6110959 | SAMEA8426056 |
| A_CC18_T_015 | A_CC18 | 2019 | Setting 4 | CC | Throat swab | 3 | EMM4.0 | 39 | ERS6110960 | SAMEA8426057 |
| A_CC22_T_008 | A_CC22 | 2019 | Setting 4 | CC | Throat swab | 2 | EMM4.0 | 39 | ERS6110961 | SAMEA8426058 |
| A_CC22_T_015 | A_CC22 | 2019 | Setting 4 | CC | Throat swab | 3 | EMM4.0 | 39 | ERS6110962 | SAMEA8426059 |
| A_CC27_T_008 | A_CC27 | 2019 | Setting 4 | CC | Throat swab | 2 | EMM4.0 | 39 | ERS6110963 | SAMEA8426060 |
| A_CC27_T_015 | A_CC27 | 2019 | Setting 4 | CC | Throat swab | 3 | EMM4.0 | 39 | ERS6110964 | SAMEA8426061 |
| A_CC28_T_008 | A_CC28 | 2019 | Setting 4 | CC | Throat swab | 2 | EMM4.0 | 39 | ERS6110965 | SAMEA8426062 |
| A_CC28_T_015 | A_CC28 | 2019 | Setting 4 | CC | Throat swab | 3 | EMM4.0 | 39 | ERS6110966 | SAMEA8426063 |
| A_CC31_T_001 | A_CC31 | 2019 | Setting 4 | CC | Throat swab | 1 | EMM89.0 | 101 | ERS6110967 | SAMEA8426064 |
| B_CC05_T_008 | B_CC05 | 2019 | Setting 5 | CC | Throat swab | 3 | EMM12.0 | 36 | ERS6110968 | SAMEA8426065 |
| B_CC05_T_046 | B_CC05 | 2019 | Setting 5 | CC | Throat swab | 4 | EMM12.0 | 36 | ERS6110969 | SAMEA8426066 |
| B_CC21_T_046 | B_CC21 | 2019 | Setting 5 | CC | Throat swab | 4 | EMM2.0 | 55 | ERS6110970 | SAMEA8426067 |
| B_CC22_T_008 | B_CC22 | 2019 | Setting 5 | CC | Throat swab | 3 | EMM4.0 | 39 | ERS6110971 | SAMEA8426068 |
| B_CC22_T_046 | B_CC22 | 2019 | Setting 5 | CC | Throat swab | 4 | EMM4.0 | 39 | ERS6110972 | SAMEA8426069 |
| B_CC34_T_001 | B_CC34 | 2019 | Setting 5 | CC | Throat swab | 2 | EMM4.0 | 39 | ERS6110973 | SAMEA8426070 |
| B_CC34_T_008 | B_CC34 | 2019 | Setting 5 | CC | Throat swab | 3 | EMM4.0 | 39 | ERS6110974 | SAMEA8426071 |
| B_CC41_T_001 | B_CC41 | 2019 | Setting 5 | CC | Throat swab | 2 | EMM4.0 | 39 | ERS6110975 | SAMEA8426072 |
| B_CC44_T_001 | B_CC44 | 2019 | Setting 5 | CC | Throat swab | 2 | EMM4.0 | 39 | ERS6110976 | SAMEA8426073 |
| B_CC44_T_008 | B_CC44 | 2019 | Setting 5 | CC | Throat swab | 3 | EMM4.0 | 39 | ERS6110977 | SAMEA8426074 |
| B_CC44_T_046 | B_CC44 | 2019 | Setting 5 | CC | Throat swab | 4 | EMM4.0 | 39 | ERS6110978 | SAMEA8426075 |
| B_CC49_T_046 | B_CC49 | 2019 | Setting 5 | CC | Throat swab | 4 | EMM4.0 | 39 | ERS6110979 | SAMEA8426076 |

| | | | | | | | | | | |
|---|---|---|---|---|---|---|---|---|---|---|
| B_E4_S_001 | B_E4 | 2019 | Setting 5 | Air | Settle plate | 2 | EMM4.0 | 39 | ERS6110980 | SAMEA8426077 |
| B_E4_S_008 | B_E4 | 2019 | Setting 5 | Air | Settle plate | 3 | EMM4.0 | 39 | ERS6110981 | SAMEA8426078 |

| | | | | | | | | | | |
|---|---|---|---|---|---|---|---|---|---|---|
| C_C1_T_010 | C_C1 | 2019 | Setting 6 | Case | Throat swab | 3 | EMM3.93 | 315 | ERS6110982 | SAMEA8426079 |
| C_C1_T_046 | C_C1 | 2019 | Setting 6 | Case | Throat swab | 4 | EMM3.93 | 315 | ERS6110983 | SAMEA8426080 |
| C_C2_T_010 | C_C2 | 2019 | Setting 6 | Case | Throat swab | 3 | EMM3.93 | 315 | ERS6110984 | SAMEA8426081 |
| C_C1_T_007 | C_C1 | 2019 | Setting 6 | Case | Throat swab | 2 | EMM3.93 | 315 | ERS6110985 | SAMEA8426082 |
| C_CC02_T_007 | C_CC02 | 2019 | Setting 6 | CC | Throat swab | 2 | EMM3.93 | 315 | ERS6110986 | SAMEA8426083 |
| C_CC07_T_007 | C_CC07 | 2019 | Setting 6 | CC | Throat swab | 2 | EMM4.0 | 39 | ERS6110987 | SAMEA8426084 |
| C_CC09_C_007 | C_CC09 | 2019 | Setting 6 | CC | Cough plate | 2 | EMM3.93 | 315 | ERS6110988 | SAMEA8426085 |
| C_CC09_H_010 | C_CC09 | 2019 | Setting 6 | CC | Hand swab | 3 | EMM3.93 | 315 | ERS6110989 | SAMEA8426086 |
| C_CC09_T_007 | C_CC09 | 2019 | Setting 6 | CC | Throat swab | 2 | EMM3.93 | 315 | ERS6110990 | SAMEA8426087 |
| C_CC13_T_007 | C_CC13 | 2019 | Setting 6 | CC | Throat swab | 2 | EMM3.93 | 315 | ERS6110991 | SAMEA8426088 |
| C_CC13_T_046 | C_CC13 | 2019 | Setting 6 | CC | Throat swab | 4 | EMM3.93 | 315 | ERS6110992 | SAMEA8426089 |
| C_CC14_C_001 | C_CC14 | 2019 | Setting 6 | CC | Cough plate | 1 | EMM3.93 | 315 | ERS6110993 | SAMEA8426090 |
| C_CC14_T_001 | C_CC14 | 2019 | Setting 6 | CC | Throat swab | 1 | EMM3.93 | 315 | ERS6110994 | SAMEA8426091 |
| C_CC15_C_007 | C_CC15 | 2019 | Setting 6 | CC | Cough plate | 2 | EMM3.93 | 315 | ERS6110995 | SAMEA8426092 |
| C_CC15_H_010 | C_CC15 | 2019 | Setting 6 | CC | Hand swab | 3 | EMM3.93 | 315 | ERS6110996 | SAMEA8426093 |
| C_CC15_T_007 | C_CC15 | 2019 | Setting 6 | CC | Throat swab | 2 | EMM3.93 | 315 | ERS6110997 | SAMEA8426094 |
| C_CC15_T_010 | C_CC15 | 2019 | Setting 6 | CC | Throat swab | 3 | EMM3.93 | 315 | ERS6110998 | SAMEA8426095 |
| C_CC15_T_046 | C_CC15 | 2019 | Setting 6 | CC | Throat swab | 4 | EMM3.93 | 315 | ERS6110999 | SAMEA8426096 |
| C_CC16_T_010 | C_CC16 | 2019 | Setting 6 | CC | Throat swab | 3 | EMM3.93 | 315 | ERS6111000 | SAMEA8426097 |
| C_CC21_T_010 | C_CC21 | 2019 | Setting 6 | CC | Throat swab | 3 | EMM3.93 | 315 | ERS6111001 | SAMEA8426098 |
| C_CC22_T_001 | C_CC22 | 2019 | Setting 6 | CC | Throat swab | 1 | EMM3.93 | 315 | ERS6111002 | SAMEA8426099 |
| C_CC22_T_007 | C_CC22 | 2019 | Setting 6 | CC | Throat swab | 2 | EMM3.93 | 315 | ERS6111003 | SAMEA8426100 |
| C_CC24_T_007 | C_CC24 | 2019 | Setting 6 | CC | Throat swab | 2 | EMM3.93 | 315 | ERS6111004 | SAMEA8426101 |
| C_CC28_T_001 | C_CC28 | 2019 | Setting 6 | CC | Throat swab | 1 | EMM4.0 | 39 | ERS6111005 | SAMEA8426102 |
| C_CC28_T_007 | C_CC28 | 2019 | Setting 6 | CC | Throat swab | 2 | EMM4.0 | 39 | ERS6111006 | SAMEA8426103 |
| C_CC34_T_010 | C_CC34 | 2019 | Setting 6 | CC | Throat swab | 3 | EMM4.0 | 39 | ERS6111007 | SAMEA8426104 |
| C_CC38_C_010 | C_CC38 | 2019 | Setting 6 | CC | Cough plate | 3 | EMM3.93 | 315 | ERS6111008 | SAMEA8426105 |
| C_CC38_T_001 | C_CC38 | 2019 | Setting 6 | CC | Throat swab | 1 | EMM3.93 | 315 | ERS6111009 | SAMEA8426106 |
| C_CC38_T_007 | C_CC38 | 2019 | Setting 6 | CC | Throat swab | 2 | EMM3.93 | 315 | ERS6111010 | SAMEA8426107 |

| | | | | | | | | | | |
|---|---|---|---|---|---|---|---|---|---|---|
| C_CC38_T_010 | C_CC38 | 2019 | Setting 6 | CC | Throat swab | 3 | EMM3.93 | 315 | ERS6111011 | SAMEA8426108 |
| C_CC39_T_007 | C_CC39 | 2019 | Setting 6 | CC | Throat swab | 2 | EMM3.93 | 315 | ERS6111012 | SAMEA8426109 |
| C_CC42_C_001 | C_CC42 | 2019 | Setting 6 | CC | Cough plate | 1 | EMM3.93 | 315 | ERS6111013 | SAMEA8426110 |
| C_CC42_C_007 | C_CC42 | 2019 | Setting 6 | CC | Cough plate | 2 | EMM3.93 | 315 | ERS6111014 | SAMEA8426111 |
| C_CC42_C_010 | C_CC42 | 2019 | Setting 6 | CC | Cough plate | 3 | EMM3.93 | 315 | ERS6111015 | SAMEA8426112 |
| C_CC42_H_001 | C_CC42 | 2019 | Setting 6 | CC | Hand swab | 1 | EMM3.93 | 315 | ERS6111016 | SAMEA8426113 |
| C_CC42_H_007 | C_CC42 | 2019 | Setting 6 | CC | Hand swab | 2 | EMM3.93 | 315 | ERS6111017 | SAMEA8426114 |
| C_CC42_T_007 | C_CC42 | 2019 | Setting 6 | CC | Throat swab | 2 | EMM3.93 | 315 | ERS6111018 | SAMEA8426115 |
| C_CC42_T_010 | C_CC42 | 2019 | Setting 6 | CC | Throat swab | 3 | EMM3.93 | 315 | ERS6111019 | SAMEA8426116 |
| C_CC51_T_010 | C_CC51 | 2019 | Setting 6 | CC | Throat swab | 3 | EMM1.0 | 28 | ERS6111020 | SAMEA8426117 |
| C_CC55_T_001 | C_CC55 | 2019 | Setting 6 | CC | Throat swab | 1 | EMM3.93 | 315 | ERS6111021 | SAMEA8426118 |
| C_CC55_T_010 | C_CC55 | 2019 | Setting 6 | CC | Throat swab | 3 | EMM3.143 | 315 | ERS6111022 | SAMEA8426119 |
| C_CC58_T_001 | C_CC58 | 2019 | Setting 6 | CC | Throat swab | 1 | EMM3.93 | 315 | ERS6111023 | SAMEA8426120 |
| C_CC58_T_046 | C_CC58 | 2019 | Setting 6 | CC | Throat swab | 4 | EMM3.93 | 315 | ERS6111024 | SAMEA8426121 |
| C_E1_S_001 | C_E1 | 2019 | Setting 6 | Air | Settle plate | 1 | EMM3.93 | 315 | ERS6111025 | SAMEA8426122 |
| C_E1_S_007 | C_E1 | 2019 | Setting 6 | Air | Settle plate | 2 | EMM3.93 | 315 | ERS6111026 | SAMEA8426123 |
| C_E1_S_010 | C_E1 | 2019 | Setting 6 | Air | Settle plate | 3 | EMM3.93 | 315 | ERS6111027 | SAMEA8426124 |
| C_E3_S_001 | C_E3 | 2019 | Setting 6 | Air | Settle plate | 1 | EMM3.93 | 315 | ERS6111028 | SAMEA8426125 |
| C_E4_S_007 | C_E4 | 2019 | Setting 6 | Air | Settle plate | 2 | EMM3.93 | 315 | ERS6111029 | SAMEA8426126 |
| C_E4_S_010 | C_E4 | 2019 | Setting 6 | Air | Settle plate | 3 | EMM3.93 | 315 | ERS6111030 | SAMEA8426127 |
| C_H2_T_010 | C_H2 | 2019 | Setting 6 | HHC | Throat swab | 3 | EMM3.93 | 315 | ERS6111031 | SAMEA8426128 |
| A_C1_T_W | A_C1 | 2018 | Setting 1 | Case | Throat swab | 0 | EMM6.0 | 382 | ERS6111032 | SAMEA8426129 |
| A_CC00_T_W1 | A_CC00 | 2018 | Setting 1 | CC | Throat swab | 1 | EMM6.0 | 382 | ERS6111033 | SAMEA8426130 |
| A_CC00_T_W3 | A_CC00 | 2018 | Setting 1 | CC | Throat swab | 3 | EMM6.0 | 382 | ERS6111034 | SAMEA8426131 |
| A_CC11_T_W1 | A_CC11 | 2018 | Setting 1 | CC | Throat swab | 1 | EMM6.0 | 382 | ERS6111035 | SAMEA8426132 |
| A_CC11_T_W2 | A_CC11 | 2018 | Setting 1 | CC | Throat swab | 2 | EMM6.0 | 382 | ERS6111036 | SAMEA8426133 |
| A_CC04_T_W2 | A_CC04 | 2018 | Setting 1 | CC | Throat swab | 2 | EMM6.0 | 382 | ERS6111037 | SAMEA8426134 |
| A_CC04_T_W3 | A_CC04 | 2018 | Setting 1 | CC | Throat swab | 3 | EMM6.0 | 382 | ERS6111038 | SAMEA8426135 |
| A_CC08_T_W3 | A_CC08 | 2018 | Setting 1 | CC | Throat swab | 3 | EMM6.0 | 382 | ERS6111039 | SAMEA8426136 |
| A_CC09_T_W3 | A_CC09 | 2018 | Setting 1 | CC | Throat swab | 3 | EMM6.0 | 382 | ERS6111040 | SAMEA8426137 |
| A_CC09_T_W4 | A_CC09 | 2018 | Setting 1 | CC | Throat swab | 4 | EMM6.0 | 382 | ERS6111041 | SAMEA8426138 |
| A_CC10_T_W1 | A_CC10 | 2018 | Setting 1 | CC | Throat swab | 1 | EMM6.9 | 382 | ERS6111042 | SAMEA8426139 |
| A_CC13_T_W2 | A_CC13 | 2018 | Setting 1 | CC | Throat swab | 2 | EMM6.0 | 382 | ERS6111043 | SAMEA8426140 |

| | | | | | | | | | | |
|---|---|---|---|---|---|---|---|---|---|---|
| A_CC13_T_W3 | A_CC13 | 2018 | Setting 1 | CC | Throat swab | 3 | EMM6.0 | 382 | ERS6111044 | SAMEA8426141 |
| A_CC13_T_W4 | A_CC13 | 2018 | Setting 1 | CC | Throat swab | 4 | EMM6.0 | 382 | ERS6111045 | SAMEA8426142 |
| A_CC14_T_W3 | A_CC14 | 2018 | Setting 1 | CC | Throat swab | 3 | EMM6.0 | 382 | ERS6111046 | SAMEA8426143 |
| A_CC15_T_W2 | A_CC15 | 2018 | Setting 1 | CC | Throat swab | 2 | EMM6.0 | 382 | ERS6111047 | SAMEA8426144 |
| A_CC15_T_W3 | A_CC15 | 2018 | Setting 1 | CC | Throat swab | 3 | EMM6.0 | 382 | ERS6111048 | SAMEA8426145 |
| B_C1_C_W3 | B_C1 | 2018 | Setting 2 | Case | Cough plate | 3 | EMM1.0 | 28 | ERS6111049 | SAMEA8426146 |
| B_C1_H_W3 | B_C1 | 2018 | Setting 2 | Case | Hand swab | 3 | EMM1.0 | 28 | ERS6111050 | SAMEA8426147 |
| B_C1_T_W3 | B_C1 | 2018 | Setting 2 | Case | Throat swab | 3 | EMM1.0 | 28 | ERS6111051 | SAMEA8426148 |
| B_C2_T_W3 | B_C1 | 2018 | Setting 2 | Case | Throat swab | 3 | EMM1.0 | 28 | ERS6111052 | SAMEA8426149 |
| B_C1_HH_W1 | B_C1 | 2018 | Setting 2 | HHC | Household | 1 | EMM1.0 | 28 | ERS6111053 | SAMEA8426150 |
| B_C2_HH_W1 | B_C2 | 2018 | Setting 2 | HHC | Household | 1 | EMM1.0 | 28 | ERS6111054 | SAMEA8426151 |
| B_C1_HH_W2 | B_C1 | 2018 | Setting 2 | HHC | Household | 2 | EMM1.0 | 28 | ERS6111055 | SAMEA8426152 |
| B_C2_HH_W2 | B_C2 | 2018 | Setting 2 | HHC | Household | 2 | EMM1.0 | 28 | ERS6111056 | SAMEA8426153 |
| B_C2_HH_W3 | B_C2 | 2018 | Setting 2 | HHC | Household | 3 | EMM1.0 | 28 | ERS6111057 | SAMEA8426154 |
| B_CC01_T_W2 | B_CC01 | 2018 | Setting 2 | CC | Throat swab | 2 | EMM1.0 | 28 | ERS6111058 | SAMEA8426155 |
| B_CC04_T_W2 | B_CC04 | 2018 | Setting 2 | CC | Throat swab | 2 | EMM1.0 | 28 | ERS6111059 | SAMEA8426156 |
| B_CC04_T_W3 | B_CC04 | 2018 | Setting 2 | CC | Throat swab | 3 | EMM1.0 | 28 | ERS6111060 | SAMEA8426157 |
| B_CC06_T_W2 | B_CC06 | 2018 | Setting 2 | CC | Throat swab | 2 | EMM1.0 | 28 | ERS6111061 | SAMEA8426158 |
| B_CC06_T_W3 | B_CC06 | 2018 | Setting 2 | CC | Throat swab | 3 | EMM1.0 | 28 | ERS6111062 | SAMEA8426159 |
| B_CC08_T_W2 | B_CC08 | 2018 | Setting 2 | CC | Throat swab | 2 | EMM1.0 | 28 | ERS6111063 | SAMEA8426160 |
| B_CC08_T_W3 | B_CC08 | 2018 | Setting 2 | CC | Throat swab | 3 | EMM1.0 | 28 | ERS6111064 | SAMEA8426161 |
| B_CC09_T_W3 | B_CC09 | 2018 | Setting 2 | CC | Throat swab | 3 | EMM1.0 | 28 | ERS6111065 | SAMEA8426162 |
| B_CC10_T_W2 | B_CC10 | 2018 | Setting 2 | CC | Throat swab | 2 | EMM1.0 | 28 | ERS6111066 | SAMEA8426163 |
| B_CC10_T_W3 | B_CC10 | 2018 | Setting 2 | CC | Throat swab | 3 | EMM1.0 | 28 | ERS6111067 | SAMEA8426164 |
| B_CC11_T_W2 | B_CC11 | 2018 | Setting 2 | CC | Throat swab | 2 | EMM12.0 | 36 | ERS6111068 | SAMEA8426165 |
| B_CC11_T_W3 | B_CC11 | 2018 | Setting 2 | CC | Throat swab | 3 | EMM12.0 | 36 | ERS6111069 | SAMEA8426166 |
| B_CC14_T_W2 | B_CC14 | 2018 | Setting 2 | CC | Throat swab | 2 | EMM1.0 | 28 | ERS6111070 | SAMEA8426167 |
| B_CC14_T_W3 | B_CC14 | 2018 | Setting 2 | CC | Throat swab | 3 | EMM1.0 | 28 | ERS6111071 | SAMEA8426168 |
| B_CC19_T_W3 | B_CC19 | 2018 | Setting 2 | CC | Throat swab | 3 | EMM6.0 | 382 | ERS6111072 | SAMEA8426169 |
| B_E1_Ty_W | B_E1 | 2018 | Setting 2 | Toy | Toys | 1 | EMM1.0 | 28 | ERS6111073 | SAMEA8426170 |
| B_E2_Ty_W | B_E2 | 2018 | Setting 2 | Toy | Toys | 1 | EMM1.0 | 28 | ERS6111074 | SAMEA8426171 |
| B_E3_Ty_W | B_E3 | 2018 | Setting 2 | Toy | Toys | 1 | EMM1.0 | 28 | ERS6111075 | SAMEA8426172 |
| B_E4_Ty_W | B_E4 | 2018 | Setting 2 | Toy | Toys | 1 | EMM1.0 | 28 | ERS6111076 | SAMEA8426173 |

| | | | | | | | | | | |
|---|---|---|---|---|---|---|---|---|---|---|
| B_E5_Ty_W | B_E5 | 2018 | Setting 2 | Toy | Toys | 1 | EMM1.0 | 28 | ERS6111077 | SAMEA8426174 |
| BR_CC12_T_W2 | BR_CC12 | 2018 | Setting 3 | CC | Throat swab | 2 | EMM1.0 | 28 | ERS6111078 | SAMEA8426175 |
| BR_CC12_T_W4 | BR_CC12 | 2018 | Setting 3 | CC | Throat swab | 4 | EMM1.0 | 28 | ERS6111079 | SAMEA8426176 |
| BR_CC17_T_W1 | BR_CC17 | 2018 | Setting 3 | CC | Throat swab | 1 | EMM1.0 | 28 | ERS6111080 | SAMEA8426177 |
| BR_CC17_T_W1_2 | BR_CC17 | 2018 | Setting 3 | CC | Throat swab | 2 | EMM1.0 | 28 | ERS6111081 | SAMEA8426178 |
| BR_CC17_T_W2 | BR_CC17 | 2018 | Setting 3 | CC | Throat swab | 2 | EMM1.0 | 28 | ERS6111082 | SAMEA8426179 |
| BR_CC18_T_W2 | BR_CC18 | 2018 | Setting 3 | CC | Throat swab | 2 | EMM1.0 | 28 | ERS6111083 | SAMEA8426180 |
| BR_CC19_T_W2 | BR_CC19 | 2018 | Setting 3 | CC | Throat swab | 2 | EMM1.0 | 28 | ERS6111084 | SAMEA8426181 |
| BR_CC19_T_W2_2 | BR_CC19 | 2018 | Setting 3 | CC | Throat swab | 3 | EMM1.0 | 28 | ERS6111085 | SAMEA8426182 |
| BR_CC20_T_W2 | BR_CC20 | 2018 | Setting 3 | CC | Throat swab | 2 | EMM1.0 | 28 | ERS6111086 | SAMEA8426183 |
| BR_CC21_T_W2 | BR_CC21 | 2018 | Setting 3 | CC | Throat swab | 2 | EMM1.0 | 28 | ERS6111087 | SAMEA8426184 |

*Genomes uploaded to ENA, project PRJEB43915. Timepoints and samples listed in supplementary tables 3-5.

**Supplementary Table 9. Genomes from Chalker *et al* (reference 3) used for analysis**

| Reference | emm | MLST |
|---|---|---|
| ERR1359668 | EMM1.0 | 28 |
| ERR1359632 | EMM1.0 | 28 |
| ERR1359833 | EMM1.0 | 28 |
| ERR1359407 | EMM1.0 | 28 |
| ERR1359854 | EMM1.0 | 28 |
| ERR1359412 | EMM1.0 | 28 |
| ERR1359400 | EMM1.0 | 28 |
| ERR1359441 | EMM1.0 | 28 |
| ERR1359627 | EMM1.0 | 28 |
| ERR1359582 | EMM1.0 | 28 |
| ERR1359530 | EMM1.0 | 28 |
| ERR1359469 | EMM1.0 | 28 |
| ERR1359819 | EMM1.0 | 28 |
| ERR1359663 | EMM1.0 | 28 |
| ERR1359370 | EMM1.0 | 28 |
| ERR1359672 | EMM1.0 | 28 |
| ERR1359419 | EMM1.0 | 28 |
| ERR1359337 | EMM1.0 | 28 |
| ERR1359866 | EMM1.0 | 28 |
| ERR1359373 | EMM1.0 | 28 |
| ERR1359723 | EMM1.0 | 28 |
| ERR1359547 | EMM1.0 | 28 |
| ERR1359825 | EMM1.0 | 28 |
| ERR1359454 | EMM1.0 | 28 |
| ERR1359440 | EMM1.0 | 28 |
| ERR1359420 | EMM1.0 | 28 |
| ERR1359732 | EMM1.0 | 28 |
| ERR1359644 | EMM1.0 | 28 |
| ERR1359427 | EMM1.0 | 28 |
| ERR1359613 | EMM1.0 | 28 |
| ERR1359670 | EMM1.0 | 28 |
| ERR1359402 | EMM1.0 | 28 |
| ERR1359385 | EMM1.0 | 28 |

| Reference | emm | MLST |
|---|---|---|
| ERR1359861 | EMM3.93 | 315 |
| ERR1359681 | EMM3.93 | 315 |
| ERR1359480 | EMM3.93 | 315 |
| ERR1359372 | EMM3.93 | 315 |
| ERR1359661 | EMM3.93 | 315 |
| ERR1359512 | EMM3.93 | 315 |
| ERR1359363 | EMM3.93 | 315 |
| ERR1359677 | EMM3.93 | 315 |
| ERR1359532 | EMM3.93 | 315 |
| ERR1359820 | EMM3.93 | 315 |
| ERR1359796 | EMM3.93 | 315 |
| ERR1359777 | EMM3.93 | 315 |
| ERR1359858 | EMM3.93 | 315 |
| ERR1359848 | EMM3.93 | 315 |
| ERR1359759 | EMM3.93 | 315 |
| ERR1359485 | EMM3.93 | 315 |
| ERR1359393 | EMM3.93 | 315 |
| ERR1359603 | EMM3.93 | 315 |
| ERR1359657 | EMM3.93 | 315 |
| ERR1359341 | EMM3.93 | 315 |
| ERR1359787 | EMM3.93 | 315 |
| ERR1359699 | EMM3.93 | 315 |
| ERR1359708 | EMM3.93 | 315 |
| ERR1359862 | EMM3.93 | 315 |
| ERR1359731 | EMM3.93 | 315 |
| ERR1359502 | EMM3.93 | 315 |
| ERR1359763 | EMM3.93 | 315 |
| ERR1359601 | EMM3.93 | 315 |

| Reference | emm | MLST |
|---|---|---|
| ERR1359656 | EMM4.0 | 39 |
| ERR1359513 | EMM4.0 | 39 |
| ERR1359639 | EMM4.0 | 39 |
| ERR1359353 | EMM4.0 | 39 |
| ERR1359593 | EMM4.0 | 39 |
| ERR1359508 | EMM4.0 | 39 |
| ERR1359808 | EMM4.0 | 39 |
| ERR1359491 | EMM4.0 | 39 |
| ERR1359749 | EMM4.0 | 39 |
| ERR1359783 | EMM4.0 | 39 |
| ERR1359660 | EMM4.0 | 39 |
| ERR1359633 | EMM4.0 | 39 |
| ERR1359450 | EMM4.0 | 39 |
| ERR1359357 | EMM4.0 | 39 |
| ERR1359543 | EMM4.0 | 39 |
| ERR1359424 | EMM4.0 | 39 |
| ERR1359678 | EMM4.0 | 39 |
| ERR1359788 | EMM4.0 | 39 |
| ERR1359446 | EMM4.0 | 39 |
| ERR1359637 | EMM4.0 | 39 |
| ERR1359576 | EMM4.0 | 39 |
| ERR1359765 | EMM4.0 | 39 |
| ERR1359339 | EMM4.0 | 39 |
| ERR1359847 | EMM4.0 | 39 |
| ERR1359662 | EMM4.0 | 39 |
| ERR1359533 | EMM4.0 | 39 |
| ERR1359643 | EMM4.0 | 39 |

| Reference | emm | MLST |
|---|---|---|
| ERR1359623 | EMM6.0 | 382 |
| ERR1359694 | EMM6.0 | 382 |
| ERR1359838 | EMM6.0 | 382 |
| ERR1359664 | EMM6.0 | 382 |
| ERR1359461 | EMM6.0 | 382 |
| ERR1359444 | EMM6.0 | 382 |
| ERR1359767 | EMM6.0 | 382 |
| ERR1359756 | EMM6.0 | 382 |
| ERR1359829 | EMM6.0 | 382 |
| ERR1359333 | EMM6.0 | 382 |
| ERR1359426 | EMM6.0 | 382 |
| ERR1359733 | EMM6.0 | 382 |
| ERR1359343 | EMM6.0 | 382 |
| ERR1359867 | EMM6.0 | 382 |
| ERR1359883 | EMM6.0 | 382 |
| ERR1359640 | EMM6.0 | 382 |
| ERR1359735 | EMM6.0 | 382 |
| ERR1359466 | EMM6.0 | 382 |
| ERR1359527 | EMM6.0 | 382 |

### Supplementary Figures

#### Supplementary Figure S1- Phylogenetic relationship between *S. pyogenes emm*6 strains from outbreak setting 1.

**A)** Maximum likelihood phylogenetic tree was constructed from 270 core SNPs (without recombination regions) extracted after mapping 17 *S. pyogenes* isolates collected from setting 1 (S1), single isolates from settings 2 and 4 (S2 and S4), and 19 *emm*6.0 scarlet fever isolates from previously published scarlet fever study in the UK (Chalker *et al*. ref 3), indicated with grey circles, to the complete *emm*6 reference strain MGAS10394 (NC_006086). Isolate source is indicated by specific shape at the end of the branch. CC (classroom contact). Settings are colour coded as indicated in the key legend. All study isolates were *emm*6.0, bar one isolate from Setting 1, *emm*6.9, which is separated from the rest of the setting 1 isolates by 40-43 SNPs. **B)** Phylogenetic tree of 16 *emm*6.0 samples from the outbreak in setting 1. Settings and sample source are colour and shape-coded as indicated in panel A. Samples of the same type (source) that are 0 SNP apart are plotted together (one on top of the other).

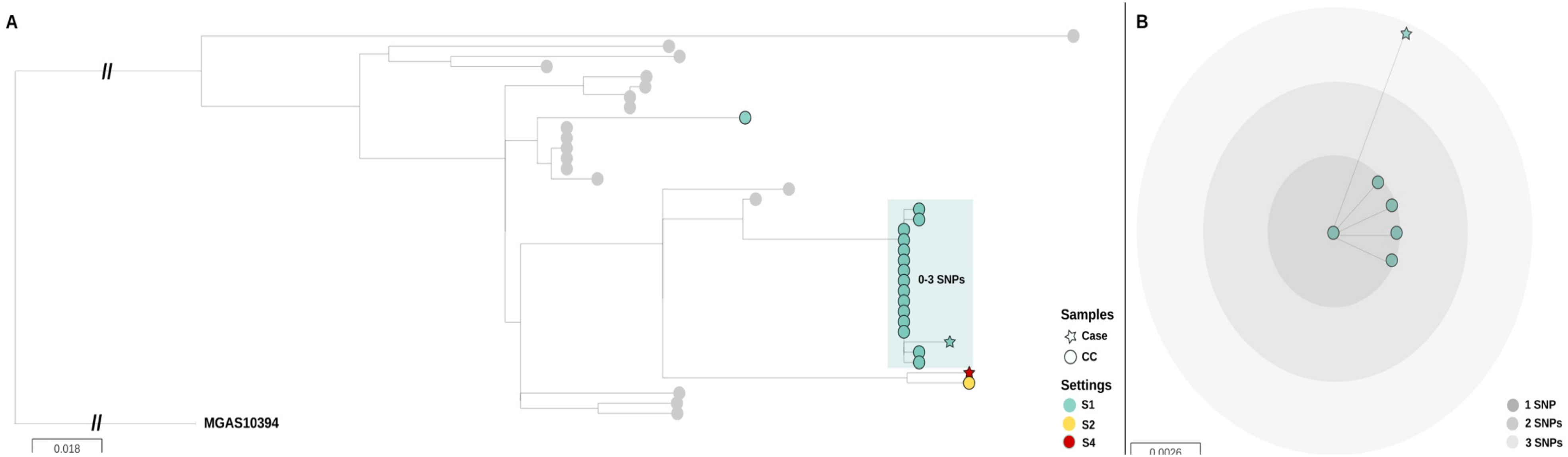

**Supplementary Figure S2**. **Phylogenetic tree of *S. pyogenes emm*1 strains from outbreak settings 2 and 3.**

**A)** Maximum likelihood phylogenetic tree constructed from 392 core SNPs (without recombination regions) extracted after mapping 37 *S. pyogenes emm*1.0 isolates collected from settings 2 and 3 (S2, S3), a single isolate from setting 6 (S6); and 33 *emm*1.0 scarlet fever isolates from previously published scarlet fever study in the UK (Chalker *et al*. ref 3), indicated with grey circles, to the complete *emm*1 reference strain MGAS5005 (CP000017). Sample source is indicated by a specific shape at the end of each branch. CC, classroom contact; HHC, household contact. Settings are colour coded as indicated in the key legend. **B)** Phylogenetic tree of 23 *emm*1 samples from setting 2. **C)** Phylogenetic tree of 10 *emm*1 samples from setting 3 and two closely related samples from setting 2. For panels B and C, settings and sample source are colour and shape-coded as indicated in panel A. Same sample types that are 0 SNP apart are plotted together; different sample types with 0 SNP apart are plotted side-by-side to facilitate visualization.

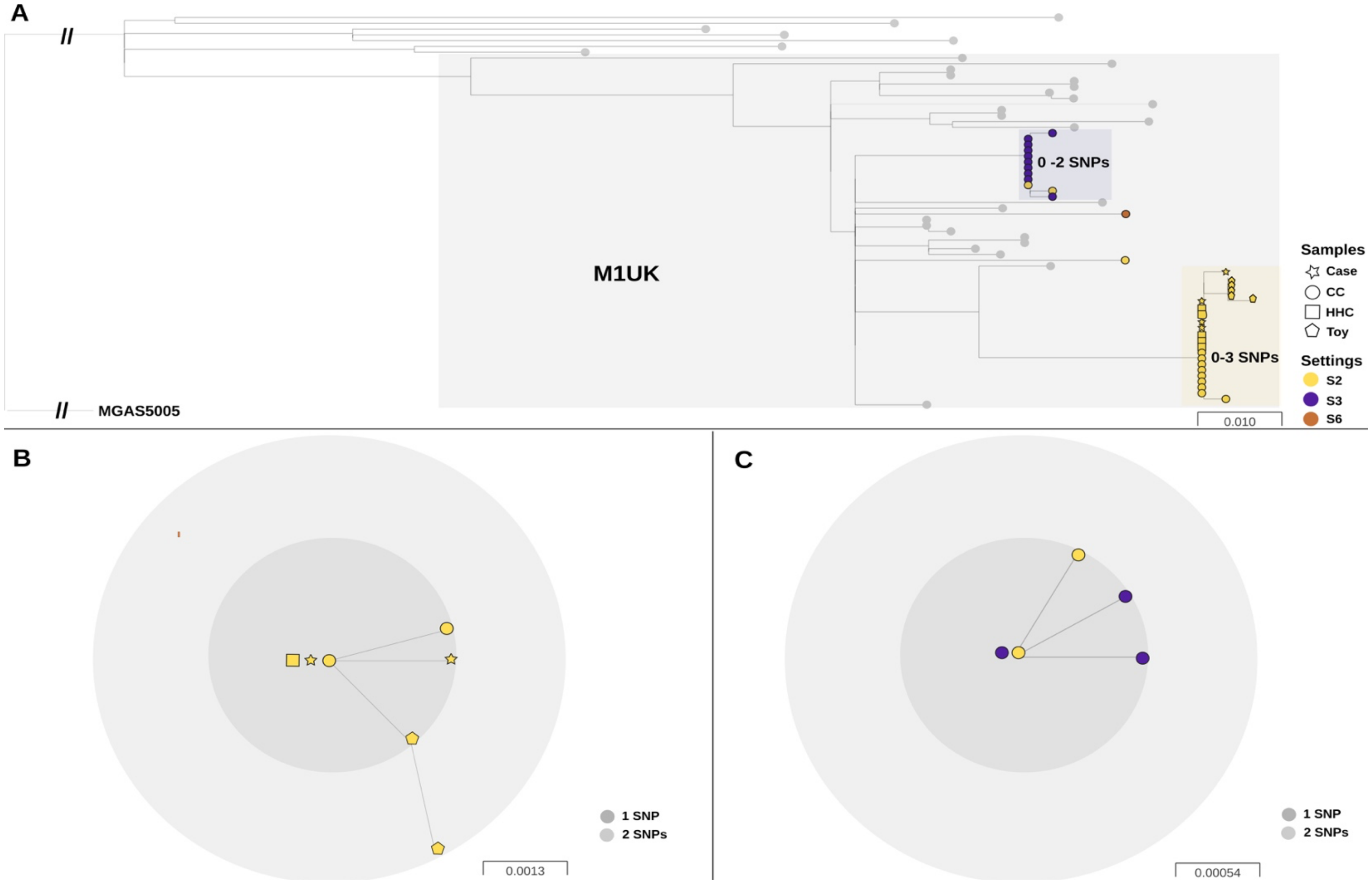

**Supplementary Figure S3**. **Phylogenetic relationship between *S. pyogenes emm*4 strains from outbreak settings 4 and 5.**

**A)** Maximum likelihood phylogenetic tree constructed from 1,005 core SNPs (without recombination regions) extracted after mapping 51 *S. pyogenes emm*4.0 isolates collected from settings 4 and 5 (S4, S5); 4 *emm*4.0 isolates from setting 6 (S6); and 27 *emm*4.0 scarlet fever isolates from previously published scarlet fever study in the UK (Chalker *et al*. ref 3), indicated with grey circles to the complete *emm*4 reference strain MGAS10750 (NC_008024). Sample source is indicated by the shape at the end of each branch. CC, classroom contact; Air, environmental settle plate. Settings are colour coded as indicated in the key legend. **B)** Phylogenetic tree of 14 *emm*4.0 isolates from setting 4 outbreak and 4 classroom contact samples from setting 6 that were closely related with up to 3SNPs difference between the isolates. Settings and sample source are colour and shape-coded as indicated in panel A. Same sample types that are 0 SNP apart are plotted together; different sample types that are 0 SNP apart are plotted side-by-side to facilitate visualization. For setting 5 and setting 4 similar visualization was not performed as no core SNPs differences were detected within the outbreak.

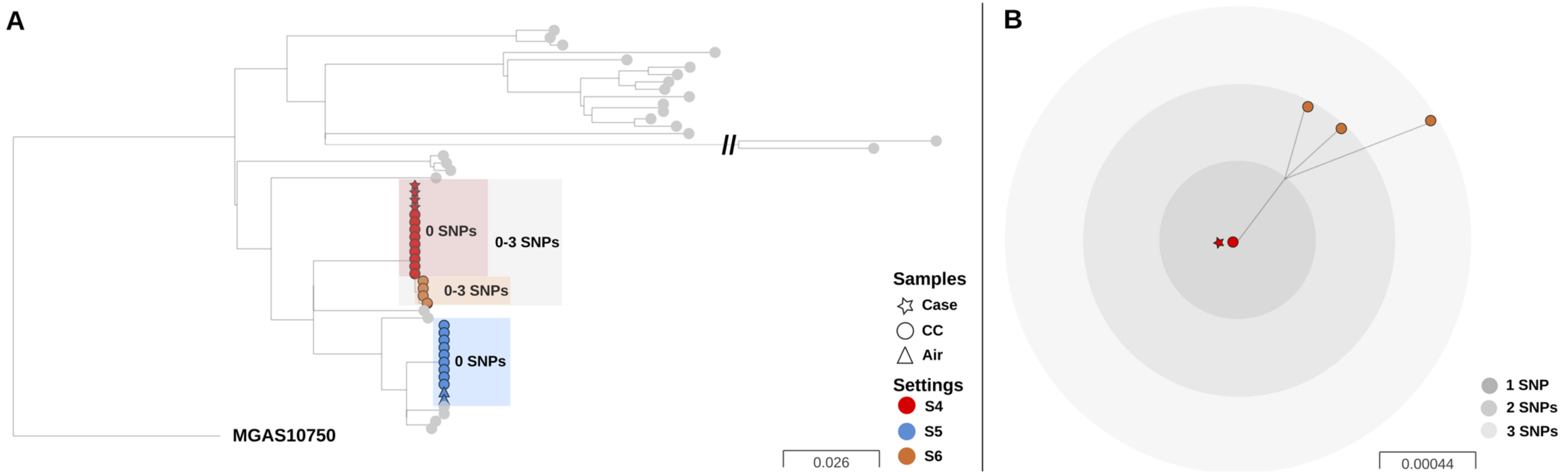

**Supplementary Figure S4**. **Phylogenetic relationship between *S. pyogenes emm*3 strains from outbreak setting 6.**

**A)** Maximum likelihood phylogenetic tree constructed from 215 core SNPs (without recombination regions) extracted after mapping 44 *S. pyogenes emm* 3.93 isolates and one 3.143 isolate collected from one setting (S6) and 28 *emm* 3.93 scarlet fever isolates from previously published scarlet fever study in the UK (Chalker *et al*. ref 3), indicated with grey circles, to the complete *emm*3 reference strain MGAS315 (NC_004070). Sample source is indicated by the shape at the end of each branch. CC, classroom contact; HHC, household contact; Air, environmental settle plate. Setting colour coded as indicated in the key legend. **B)** Phylogenetic tree of all 45 *emm*3 isolates from setting 6 outbreak. Settings and sample source are colour and shape-coded as indicated in panel A. Same sample types that are 0 SNP apart are plotted together; different sample types that are 0 SNP apart are plotted side-by-side to facilitate visualization.

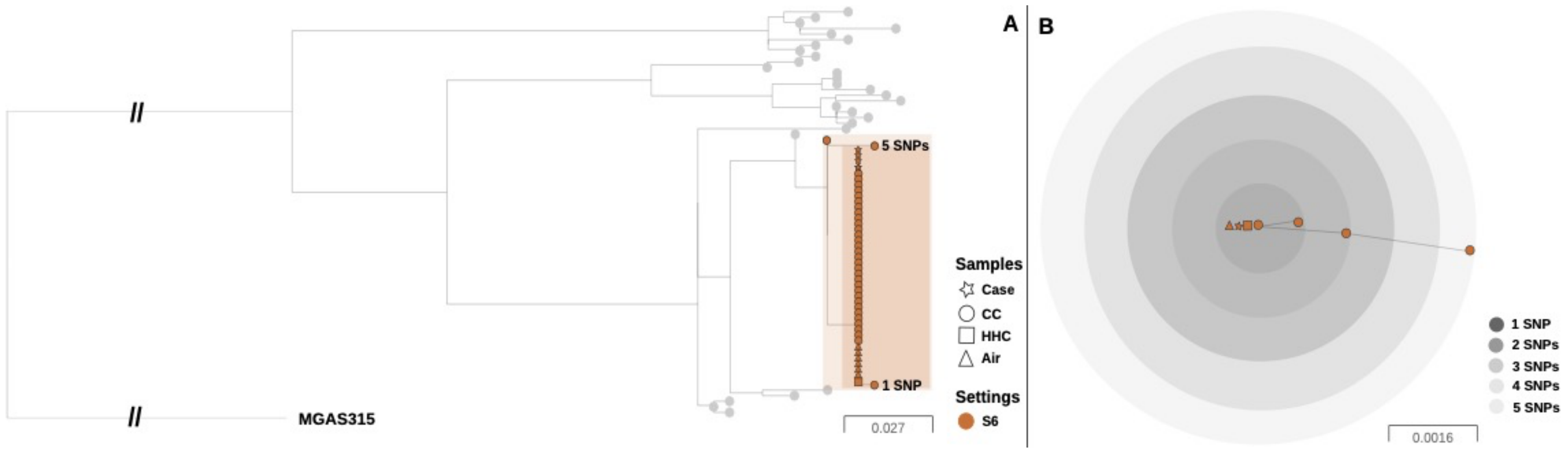

### Supplementary Methods

*Case definition.* Confirmed cases were those with a clinical diagnosis of scarlet fever by a health professional (sore throat, fever, sand-papery rash) and *Streptococcus pyogenes* grown from a throat swab. Probable cases were those with a clinical diagnosis of scarlet fever by a health professional without bacteriological confirmation. Possible cases were those reported by a reliable source (e.g. nursery or school manager), presenting with signs and symptoms consistent with scarlet fever, and a close epidemiological link e.g. household contact of a confirmed case; or attending school where there is a confirmed scarlet fever outbreak.

*Recruitment of schools.* Confirmed and probable cases of scarlet fever were identified by notifications to local Health Protection Teams. Schools and nurseries were invited to participate if they had two confirmed or probable scarlet fever cases aged 2-8 years from the same class within ten days of each other, with the most recent case arising in the preceding 48h. Locations were excluded if reported cases did not match criteria for cases, if timings did not match inclusion criteria, or if they declined.

*Bacteriology and sampling.* Swabs from participants were transported in Amies medium (Deltalab, Barcelona, Spain), then plated immediately onto Columbia Blood Agar (CBA, Oxoid, Basingstoke, UK). For cough plates, participants were encouraged to cough onto CBA held at a distance of 20cm from the mouth. CBA plates were incubated in a BSL2 laboratory at 37°C 5% $CO_2$ overnight then inspected for beta-haemolytic colonies. *S. pyogenes* was identified using Bruker MALDI-ToF Biotyper (Bruker Daltonics, Bremen, Germany). To evaluate presence of *S. pyogenes* DNA in stored culture-negative throat swabs, the *S. pyogenes* housekeeping gene *proS* was amplified from extracted DNA using primers *ProS F* 5′TGAGTTTATTATGAAAGACGGCTATAGTTTC and *ProS R* 5′-AATAGCTTCGTAAGCTTGACGATAATC and copies of *proS* were quantified by comparison with standard concentrations of a plasmid containing a single copy of *proS* as described previously (1). The lower 99% confidence interval of the geometric mean value obtained from 66 culture-positive throat samples from 2019 was used as a cut off (~ 91.6 copies/swab).

In year 1, surface samples (25cm$^2$) were obtained from frequently touched surfaces in classrooms in week 1 only (20 samples per classroom) using dry cotton swabs moistened in sterile saline. Swabs were placed into 1 ml of sterile saline, and transported to the laboratory where they were diluted using a 10 times serial dilution in sterile phosphate buffered saline to a dilution of $10^{-6}$. Duplicate 50 µl volumes of each dilution were plated on to CBA plates and incubated at 37°C in air supplemented with 5% $CO_2$ for 48h.

In year 2, air settle plates were used to detect airborne dispersal of *S. pyogenes* using CBA plates placed on horizontal surfaces that were at least 1.5m high such as shelves and cupboards. A pilot study (setting 4) revealed that plates left for 24h were overgrown and un-readable, therefore, for settings 5 and 6, settle plates were left for 2-3h while children were using the classroom. Four plates were used per classroom per time point in settings 4-6, on each of the weeks studied.

*Genomic analysis*. DNA was extracted from all cultured *S. pyogenes* isolates from overnight cultures and *emm* genotyping was performed according to the protocol of the Centers for Diseases Control and Prevention (www.cdc.gov/ncidod/biotech/strep/protocol_emmtype) followed by pair-end 150bp read length whole genome sequencing on an Illumina HiSeq 2000 platform (Illumina, USA) according to the manufacturer's protocol. In 2019 (settings 4-6) *emm*-typing was performed only from whole genome sequencing data using BLAST+ version 2.2.30 against a specific *S. pyogenes emm*-type database (https://www2.cdc.gov/vaccines/biotech/strepblast.asp). Sequence data have been submitted to the European Nucleotide Archive (ENA - www.ebi.ac.uk/ena) under the accession number PRJEB43915 (Supplementary table 8). Raw reads were trimmed using trimmomatic version 0.36 (2) with the following parameters: trimCrop=N, trimHeadCrop=N, sliding window 5:20, trim leading=3, trim trailing=4, trim min length=55, before any further downstream analysis. The comparative SNP-calling analysis was performed by mapping trimmed reads of 136 *S. pyogenes* isolates to the complete *emm*89 reference sequence H293 (HG316453.2) using Snippy v4.6.0 (https://github.com/tseemann/snippy), with a minimum coverage of 10, minimum fraction of 0.9, and minimum vcf variant call quality of 100.

Gubbins version 2.4.1 (3) was used to identify and remove recombinant regions from the resulting full genome alignment file. A maximum likelihood phylogeny was created from core SNPs using the general time-reversible (GTR) model of nucleotide substitution with the gamma distributed rate heterogeneity implemented in FastTree v2.1.10-4 (4) Phylogenetic trees were visualized using FigTree v1.4.2 (http://tree.bio.ed.ac.uk/software/figtree/) and Microreact (https://microreact.org/showcase) and edited using INKSCAPE (https://inkscape.org/pt/). Genetic diversity within *emm*-types identified as dominating in each setting (*emm 1*, *emm 3*, *emm 4* and *emm 6*) was assessed by comparison with previously sequenced isolates from an earlier UK scarlet fever study (5) (**Supplementary table 9**) and using *emm*-type specific reference genomes as follows: for *emm1,* MGAS5005 (CP000017); for *emm3,* MGAS315 (NC_004070); for *emm4*, MGAS10750 (NC_008024); for *emm6,*

MGAS10394 (NC_006086). The SNP distance matrix was obtained using snp-dist (https://github.com/tseemann/snp-dists). SNPs identified within each outbreak setting were classified as non-coding, missense or synonymous according to the location in the genome and effect on protein using Snippy. The functional effect of each amino acid substitution was predicted using PROVEAN Protein database (http://provean.jcvi.org/seq_submit.php). *emm*4 sub-lineage classification (M4 "complete" vs M4 "degraded") was performed as described by Remmington *et al.* 2021 (6), while *emm*1 sub-lineage classification (M1 vs $M1_{UK}$) was performed as described by Lynskey, Jauneikaite *et al.* 2019 (7).
